# Same pipelines, different answers: how evaluation design shapes EEG seizure prediction

**DOI:** 10.64898/2026.10.28.26366763

**Authors:** Zikang Song, Mangor Pedersen, Dion Henare

## Abstract

Seizure-prediction studies on scalp electroencephalography (EEG) often report accuracies that would be clinically useful, yet the score a model receives also depends on how its test data were separated from its training data. We kept five published pipelines fixed, from a spectral baseline (power spectral density with linear discriminant analysis, PSD+LDA) to a transformer, and changed only how they were evaluated on the CHB-MIT database. When whole subjects were held out in 20 repeated partitions, every pipeline scored close to the 0.5 reference (mean area under the ROC curve, AUC, 0.527–0.553), and the five pipelines differed less from one another than each varied between partitions. Within patients, PSD+LDA reached an AUC of 0.815 when windows were allocated at random but 0.572 when each preictal block was kept intact and later seizures were held out for testing. Tracing every window back to its recording showed that an index-based within-patient split had let raw EEG samples cross the train–test boundary in all ten eligible cases. Thresholds tuned to allow 1.67% false positives on validation data did not hold that rate on new subjects, and at those thresholds only 2–6% of preictal windows were detected. A zero-shot test of the baseline on the Siena database gave an AUC of 0.480. In this benchmark, the choice of evaluation changed the answer far more than the choice of architecture. We release the window manifests, partitions and predictions so that new models can be tested on the same terms. All results concern retrospective window classification, not continuous alarms.

## Introduction

A reliable seizure warning would give a person time to move somewhere safe or use a planned intervention before onset. For that to help, the warning has to catch most seizures without raising so many false alarms that people stop trusting it [1–3]. Most EEG studies approach this goal through a narrower task: telling selected preictal windows apart from selected interictal ones. Reported accuracies on that task are often high, but they are hard to read when a paper does not say what the training and test data shared, or what a positive decision is supposed to trigger [4, 5].

Several groups have shown how fragile such numbers can be. Shafiezadeh et al. found that strong cross-validation results did not carry over to new patients [6]. Brookshire et al. showed how segment-level leakage inflates translational EEG classifiers [7], and reviews of seizure prediction note how rarely models are tested on independent patients or external data [8, 9]. What remains open is a question that anyone reading a results table has to answer: if the models stay the same and only the evaluation changes, how far does the answer move, and what information crosses the split under each design?

The answer depends on details that rarely make it into a methods section. Neighbouring EEG windows can share raw samples, windows from the same seizure share its surrounding background activity, and different recordings can come from the same person. A pooled AUC compares preictal and interictal windows across subjects, whereas a subject-wise AUC compares them within one person. Allocating windows at random, testing on a patient’s later seizures and testing a person with no prior data are therefore different tasks, even when the classifier and labels do not change.

Access to the target patient changes the task again. Calibration and domain adaptation can improve patient-independent models once some target data are available [10–13], and Jemal et al. evaluated cross-subject prediction with domain adaptation [14]. Comparing such results with a cold-start model is fair only after accounting for target access, label definitions and window construction.

A working system also needs a threshold, and AUC says nothing about how a threshold behaves on new people. A cut-off that suppresses positive decisions on validation recordings may not do so on unseen subjects. Even a stable false-positive proportion among sparsely sampled interictal windows is not an alarm rate: that requires a temporal alarm policy, with rules for persistence, merging and refractory periods, and a denominator of monitoring time [5]. Preprocessing that looks into the future is a further obstacle to online use [15].

Here we kept five established pipelines fixed and changed only the evaluation. We tested them on subject groups they had never seen, traced every EEG window to its recording and seizure so that complete episodes could be kept together in time, and carried validation-selected thresholds unchanged to new subjects. Depending on these choices, the same pipelines looked either close to the 0.5 reference or clearly discriminative. The window manifests, partitions, archived predictions and analysis code are released so that new models can be tested on the same terms. Analyses that deliberately reuse samples, and the original index-based allocation, are kept in the supplement as diagnostics; neither isolates an effect of leakage.

## Materials and methods

### Study design and analysis hierarchy

This retrospective computational study was not preregistered. The primary analysis is the original benchmark: 20 strict subject-group partitions and five fitted pipelines. The chronological block control and the validation false-positive constraint were added afterwards, once the original results had been inspected. Their rules were written down before their outcomes were summarized, and no test outcome was used to choose an architecture, a temporal boundary or a threshold. Figure 1 shows what information each analysis could use.

**Figure 1.**
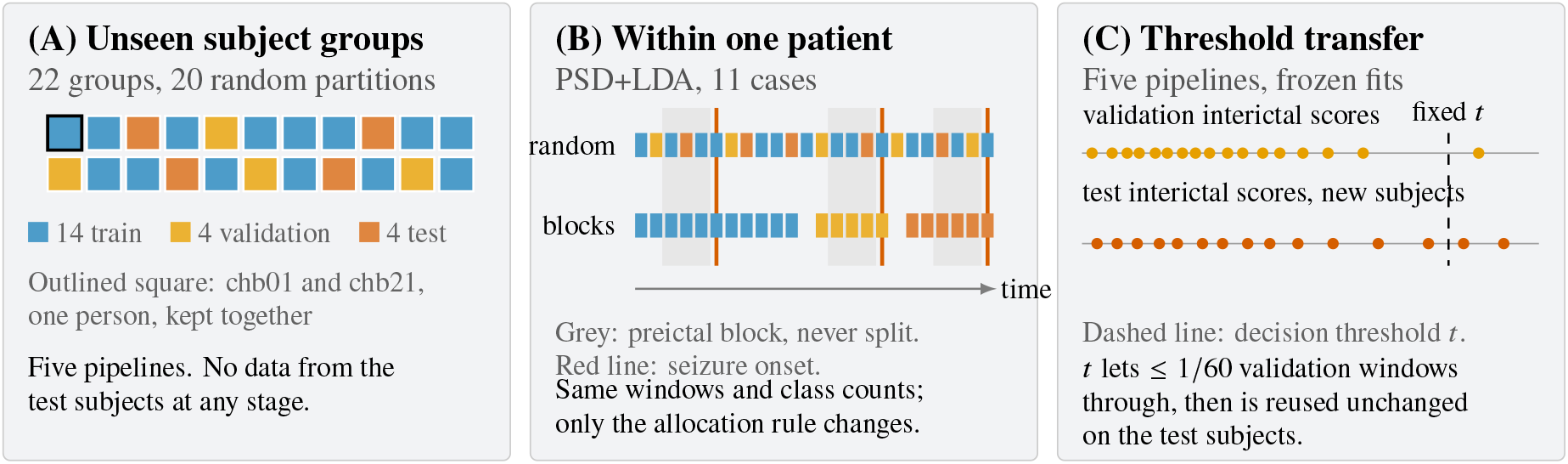
The three evaluation designs. (A) Whole subject groups are held out for cold-start testing; the two CHB-MIT folders from one person stay in the same partition. (B) Within each eligible case, the same windows are allocated either at random or in time order with every preictal block kept intact, using identical class counts. (C) A threshold is chosen from validation interictal scores alone and then applied, unchanged, to held-out subject groups. Square and window counts are schematic. All results concern retrospective windows.

### Datasets and signal preparation

We used CHB-MIT version 1.0.0 [16–19]. The original chb01–chb23 case folders contain 182 annotated seizures from 22 pediatric participants. The folders chb01 and chb21 belong to the same participant, recorded approximately 1.5 years apart, so they were treated as one subject group. The later-added chb24 retained no interictal windows under the present exclusion rules and was omitted. The common cohort contained 23 case identifiers and 22 independent subject groups.

We retained EDF recordings sampled at 256 Hz with 18 shared bipolar derivations: FP1–F7, F7–T7, T7–P7, P7–O1, FP1–F3, F3–C3, C3–P3, P3–O1, FP2–F4, F4–C4, C4–P4, P4–O2, FP2–F8, F8–T8, T8–P8, P8–O2, FZ–CZ and CZ–PZ. Legacy T3/T4/T5/T6 labels were mapped to T7/T8/P7/P8, and duplicate channel labels were removed. Recording times were reconstructed from EDF measurement dates when usable, otherwise from summary clocks with midnight rollover, and finally from the preceding recording’s end when neither was available. Each EDF was filtered separately from 0.5 to 40 Hz using MNE’s default zero-phase finite impulse response filter.

Windows lasted 20 s. A preictal window had to lie entirely 5–30 min before an annotated onset; preictal windows advanced by 10 s and so overlapped by 10 s. Interictal windows normally advanced by 300 s, with 10-s steps near a label transition according to the released preprocessing routine. Ictal periods, the final 5 min before onset and 4 h after seizure termination were excluded [1, 20]. Each retained window was z-score normalized independently for each channel, and windows with more than two channels having standard deviation below 10^−6^ were removed. No normalization statistic was estimated across windows or partitions.

The resulting data contained 17,424 windows. Preictal windows came from 69 of the 182 annotated seizures. This count uses annotation identities, including multiple memberships when preictal intervals overlap. It differs from the 60 contiguous positive-label runs in the concatenated arrays, because such runs can merge separate events or cross recording gaps. Supplementary Table S1 reports the case-level counts and eligibility.

For the secondary external control, we used Siena Scalp EEG version 1.0.0 [21, 22]. The existing adapter resampled 512-Hz recordings to 256 Hz, constructed the same bipolar montage and applied the same window definitions, exclusions, filtering and PSD extraction. Thirteen participants retained both classes, giving 5,657 windows (4,471 preictal and 1,186 interictal). PN00 was excluded because no interictal window remained. Only PSD+LDA was evaluated externally, and no Siena data were used for fitting, threshold selection or calibration.

### Models and training

The five pipelines were PSD+LDA, EEGNet, a temporal convolutional network (TCN), a one-dimensional convolutional network (1D-CNN) and EEG-Conformer. PSD+LDA used Welch spectra with 512-sample segments. Mean power in five bands (0.5–4, 4–8, 8–13, 13–30 and 30–40 Hz, endpoints inclusive) gave 90 features across 18 channels. Linear discriminant analysis used the singular-value-decomposition solver and equal class priors, with no feature selection.

EEGNet used eight temporal filters, depth multiplier 2 and a 127-sample temporal kernel [23]. The TCN used four 64-filter residual blocks, kernel size 8 and dilations 1, 2, 4 and 8 [24]. The 1D-CNN used three stages with parallel kernels of sizes 3, 5 and 7. EEG-Conformer used 40-dimensional patch embeddings, ten attention heads and six encoder blocks [25]. These deep models had 7,002, 245,122, 520,322 and 852,746 parameters, respectively. They are the released implementations; the benchmark does not include an architecture search.

Deep models used cross-entropy loss, AdamW with weight decay 10^−4^, gradient clipping at 1.0 and a weighted training sampler with replacement. EEGNet and TCN used learning rate 10^−3^ and batch size 128; 1D-CNN used 10^−4^ and 128; EEG-Conformer used 10^−4^ and 64. Training stopped after at most 100 epochs or 20 epochs without improvement in validation AUC, and the checkpoint with the highest validation AUC was retained. A reduce-on-plateau scheduler halved the learning rate after five non-improving validation epochs, down to 10^−7^. The later analyses reused these fitted weights without retraining.

### Cold-start subject-group evaluation

For each of 20 fixed seeds, the 22 subject groups were assigned to 14 training, four validation and four test groups, with the chb01/chb21 group kept intact. Depending on where that group fell, this gave 14 or 15 training case identifiers and four or five validation or test identifiers. The same seed controlled allocation and model fitting. Every group was tested at least once, with one to eight test appearances across the 20 partitions (Supplementary Table S2). No target-subject data were available for training, early stopping or threshold selection.

The primary outcome was AUC after pooling all test windows within a partition, summarized as the mean and sample standard deviation (SD) over the 20 partitions. We also calculated AUC separately for each subject group at each test appearance, averaged appearances within a group, and weighted the 22 groups equally:

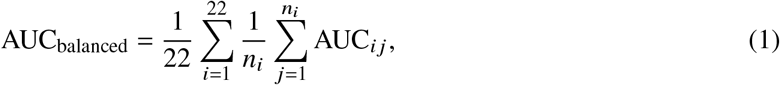

where *n*_*i*_ is the number of test appearances for group *i*. Its SD is the between-group sample SD. Pooled AUC reflects unequal numbers of windows and includes positive–negative comparisons across subjects, so it is not simply a weighted average of within-subject AUCs.

The original operating threshold maximized validation Youden’s index. When validation AUC was below 0.5, the implementation fell back to a threshold of 0.5. This happened in nine of 100 model-by-partition runs: six for PSD+LDA and one each for EEGNet, TCN and 1D-CNN. We kept this rule for the original results and report pooled window sensitivity and specificity.

### Window provenance and chronological block control

We reconstructed candidate window positions from EDF headers and the original annotations using the released preprocessing rules. Counts and label sequences matched the saved arrays directly in 22 cases. In chb17, applying the original filtered flatline rule removed seven candidate windows and recovered the saved sequence exactly. For this case, all 491 retained waveforms were compared with the saved data; in the other 22 cases, 126 representative windows were checked across up to two EDFs per case. The maximum absolute difference after normalization was 5.96 × 10^−8^. The resulting map records, for every window, its recording, sample position, reconstructed time and every seizure it is associated with.

The original within-case audit cut the concatenated window array at 60% and 80% by index. Its ten eligible cases had both classes in training, validation and test sets. We checked these boundaries for shared seizure identities and overlapping raw samples, and repeated the PSD+LDA calculation after sorting windows by reconstructed start time. The original five-model random-versus-index results are kept in the supplement, with the allocation rule stated as it was actually applied.

For the chronological block control, overlapping nominal preictal intervals were joined into indivisible blocks, which also kept windows with multiple seizure memberships together. Blocks were ordered by reconstructed time, and candidate boundaries were the midpoints between successive retained block spans. We chose the pair of boundaries closest to cumulative window fractions of 0.60 and 0.80 (minimizing the sum of squared deviations after exclusion), subject to both classes appearing in each partition. Windows had to lie at least 10 s from a boundary, and all windows from a block stayed in one partition. Boundary selection used timing and labels, never model scores. Actual partition sizes were allowed to differ from 60/20/20; training fractions ranged from 17.8% to 67.7% (Supplementary Table S3).

Eleven cases met these rules. We fitted PSD+LDA on each case’s training partition and evaluated its test partition. For comparison, 20 random allocations used the same retained windows and exactly matched the preictal and interictal counts of each blocked partition, shuffling each class separately with the fixed seed list. Random-split AUCs were averaged within each case before paired differences were summarized. We chose PSD+LDA for this control because it has fixed features and no checkpoint selection; the control was not repeated for the deep models. The data were still filtered retrospectively, so the control tests separation of evaluation periods rather than online prediction.

### Transfer of a validation false-positive constraint

For this analysis we defined an operating rule from the validation interictal scores alone. Let *N*_0_ be their number and *k* = ⌊*N*_0_/60⌋ the permitted number of false positives. After sorting these scores in descending order, the threshold was set just above the (*k* + 1)th score. With the decision rule *p ≥ t*, this gives the smallest representable threshold that meets the validation constraint, with ties handled conservatively. The target false-positive proportion of 1/60 (about 1.67%) is an illustrative operating point with no claimed clinical status.

Validation predictions were recovered from the frozen deep-model checkpoints, and PSD+LDA fits were rebuilt from the original cached features and training partitions. Before the validation scores were used, recovered test scores were checked against the archived predictions: the maximum absolute discrepancy was below 6.4× 10^−4^ across pipelines, and deep-model validation AUCs agreed with the checkpoint records within 10^−4^. Held-out results were scored with the original archived test predictions so that they remain traceable to the original benchmark. Numerical sensitivity to score recovery is reported in the supplement.

The threshold was then applied unchanged to the held-out subject groups. We report the observed test false-positive proportion, window sensitivity and the number of partitions in which the validation target was exceeded. These are decisions among sampled windows, and we do not convert nominal sampling intervals into monitoring time. A cadence-adjusted metric used in earlier analyses, and a diagnostic whose threshold was chosen on test data, are reported in the supplement and kept apart from the validation-selected policy.

### Statistical interpretation

The repeated subject partitions reuse participants and training data. Their SDs describe variation across partitions and do not provide 20 independent estimates of population performance, so we do not report ordinary *t* confidence intervals across splits. The subject-balanced SD describes variation among the 22 group means, whose fitted models are also shared. The chronological block control uses the case as the descriptive unit (*n* = 11) and is not combined with the ten-case index audit.

The original ten-case audit used two-sided Wilcoxon signed-rank tests, pairing each index-split AUC with that case’s mean random-split AUC, with a Bonferroni threshold of 0.01 for the five model-wise comparisons. These exploratory tests are reported in the supplement. There was no prospective sample-size calculation and no formal test against a temporal surrogate null distribution. AUC 0.5 is a nominal ROC reference, not evidence that an estimate is statistically equivalent to chance [26]. We did not assess probability calibration, event sensitivity or continuous alarm performance.

## Results

### On unseen subjects, the five pipelines were hard to tell apart

Across the 20 partitions, mean pooled AUC ran from 0.527 (PSD+LDA and EEG-Conformer) to 0.553 (TCN), a spread of 0.026 between models. A single pipeline varied more than that from one partition to the next, with SDs of 0.06–0.08 (Table 1). Weighting the 22 subject groups equally brought the means down to 0.502–0.523. It also exposed how unevenly the models behaved across people: some subject groups scored well below 0.5 and others well above it (Figure 2B). Even the ranking depended on the summary, with TCN ahead on pooled AUC and 1D-CNN ahead on the subject-balanced mean. Neither difference supports an architecture ranking.

**Table 1.** Cold-start results. Pooled AUC, sensitivity and specificity are mean ± SD across 20 subject-group partitions. Subject-balanced AUC is mean ± between-group SD across 22 groups. Sensitivity and specificity use the original validation Youden rule, including its recorded fallback.

| Pipeline | Pooled AUC | Subject-balanced AUC | Sensitivity | Specificity |
| --- | --- | --- | --- | --- |
| PSD+LDA | 0.527 ± 0.070 | 0.502 ± 0.111 | 0.460 ± 0.202 | 0.584 ± 0.185 |
| EEGNet | 0.534 ± 0.060 | 0.519 ± 0.090 | 0.407 ± 0.214 | 0.649 ± 0.190 |
| TCN | 0.553 ± 0.070 | 0.515 ± 0.121 | 0.514 ± 0.261 | 0.555 ± 0.221 |
| 1D-CNN | 0.541 ± 0.078 | 0.523 ± 0.133 | 0.380 ± 0.236 | 0.652 ± 0.199 |
| EEG-Conformer | 0.527 ± 0.069 | 0.517 ± 0.120 | 0.462 ± 0.278 | 0.568 ± 0.235 |

AUC: area under the receiver-operating-characteristic curve; SD: standard deviation; PSD+LDA: power spectral density with linear discriminant analysis; TCN: temporal convolutional network; 1D-CNN: one-dimensional convolutional neural network.

**Figure 2.**
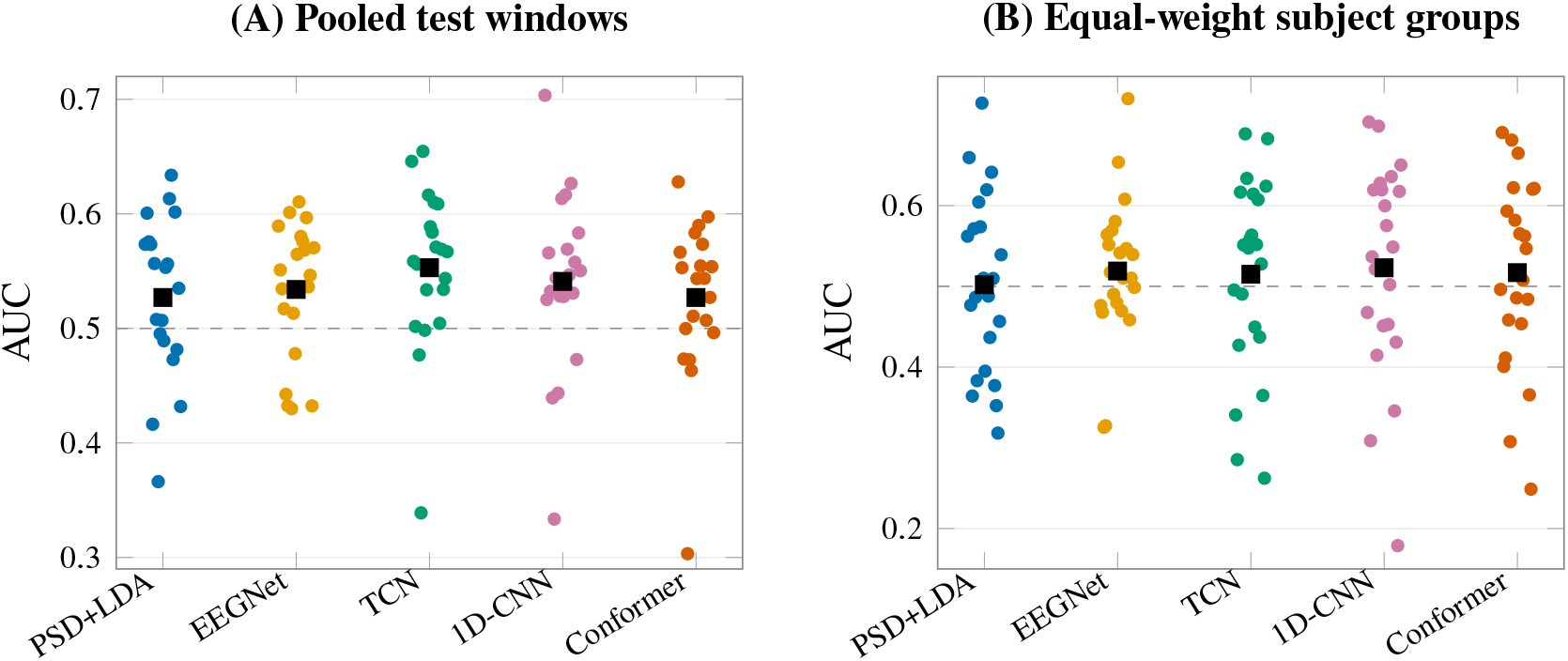
AUC on unseen subject groups. (A) Pooled AUC for each of the 20 held-out partitions. (B) AUC within each subject group, averaged over that group’s test appearances; one point per group (22 groups). Black squares are means. The dashed line marks the nominal 0.5 ROC reference. Points are jittered horizontally only to reduce overlap.

### Keeping seizure episodes together lowered within-patient AUC

Tracing windows back to their recordings changed how the original within-patient audit should be read. In all ten cases eligible for that audit, at least one seizure contributed windows to more than one partition, and at least one pair of windows shared raw samples across a boundary (16 such pairs in total). The original random-minus-index differences of 0.139–0.373 across the five pipelines therefore compare two allocation schemes that both let related windows cross the split (Supplementary Tables S4 and S5). One out-of-order window in chb03 did not change the eligible cohort or the PSD+LDA results once windows were sorted by time.

The chronological block control closed these gaps. In 11 eligible cases (11,175 windows), no seizure block and no pair of overlapping windows crossed a partition, and no window had to be dropped at a boundary. Under this design PSD+LDA reached a mean AUC of 0.572, against 0.815 when the same windows were allocated at random with identical class counts. Random allocation scored higher in all 11 cases, by 0.243 on average and by as much as 0.601 (Figure 3). The gap was largest in chb04, chb13 and chb15, where the chronological AUC fell below 0.4 while the random AUC stayed between 0.79 and 0.91. Because these 11 cases are not the original ten, the two comparisons cannot be read as a before-and-after estimate of correcting the boundaries.

**Figure 3.**
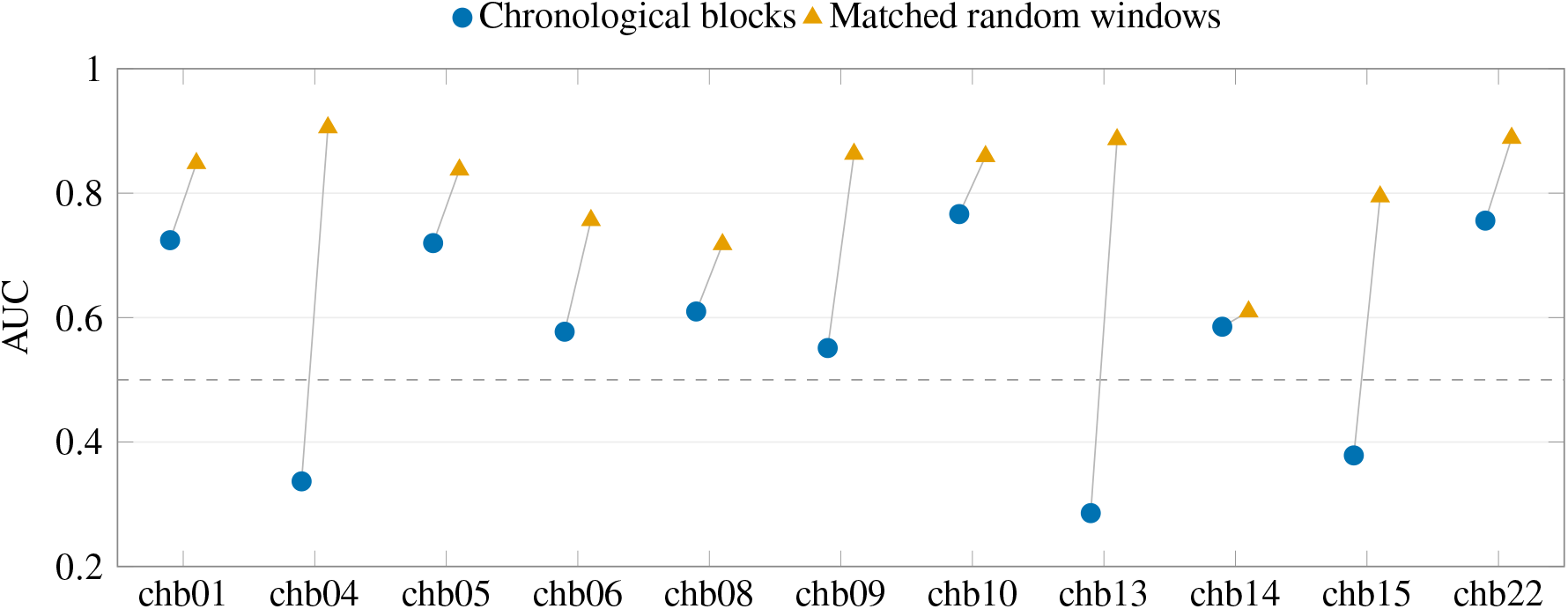
PSD+LDA within individual patients, under chronological allocation of complete preictal blocks and under matched random allocation of windows. Each line joins the two estimates for one case. Random values are means over 20 allocations with the same train/validation/test class counts as the corresponding blocked split. Across the 11 cases, AUC was 0.572 ± 0.171 for blocked and 0.815 ± 0.089 for random allocation (mean ± SD); the paired difference was 0.243 ± 0.201, ranging from 0.024 to 0.601. Changing the allocation rule changes both the sharing of temporally related windows and the time span covered by training and test data, so the gap does not identify one isolated source of optimism.

### Validation false-positive limits did not hold on unseen subjects

Under the original Youden rule, the pipelines detected about half of preictal windows or fewer, while classifying a third or more of interictal windows as positive (Table 1). We therefore asked whether a stricter threshold, chosen on validation data, would keep its false-positive rate on new people. It did not do so reliably. Thresholds that let through 1 in 60 validation interictal windows gave mean test false-positive proportions of 1.81–3.13%, and every pipeline exceeded the validation target in 6–10 of the 20 partitions (Figure 4A; Supplementary Table S6). Some partitions stayed well under the target while others exceeded it several times over, although the selection rule never changed. Sensitivity fell at the same time: on average, only 2.33–5.56% of preictal windows were detected (Figure 4B).

**Figure 4.**
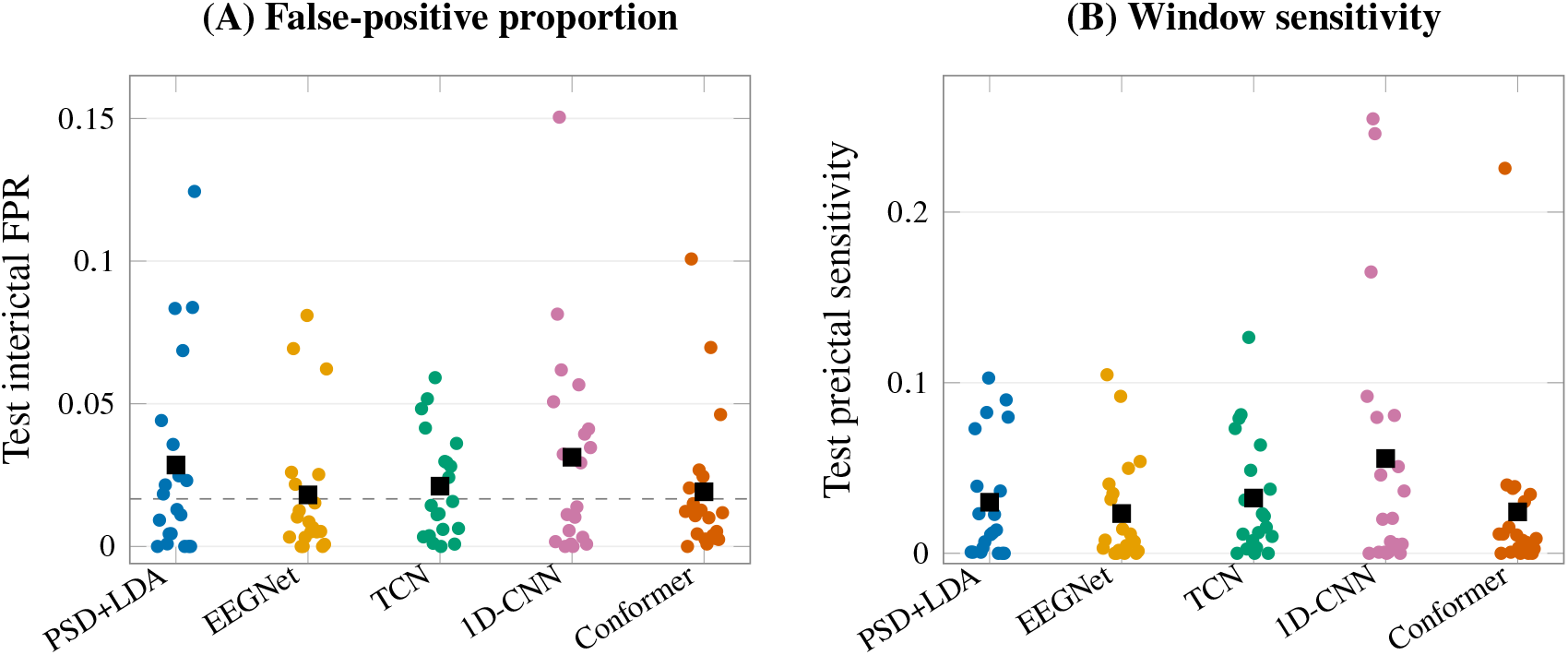
Held-out operating points after choosing each threshold so that at most 1/60 of validation interictal windows were classified as positive. (A) Test interictal false-positive proportion. The dashed line is the validation target, which is not imposed on the test set. (B) Test preictal window sensitivity. Each point is one of 20 subject-group partitions; black squares are model means. Both panels report proportions among sampled windows, without conversion to alarms or monitoring hours.

### The baseline stayed near 0.5 on a second database

Applied without any refitting to the 13 Siena participants, the 20 CHB-MIT-trained PSD+LDA fits gave a mean pooled AUC of 0.480 ± 0.022 (participant-macro AUC 0.477 ± 0.024). The CHB-MIT validation Youden thresholds detected on average 19.5% of Siena preictal windows. This is a limited check on the portability of the feature pipeline; the deep models were not run on Siena (Supplementary Table S8).

## Discussion

With the pipelines held fixed, changing the evaluation moved the answer far more than changing the architecture. On unseen subject groups, five models ranging from a 90-feature linear classifier to a transformer with more than 850,000 parameters differed by less than 0.03 in mean AUC, and all sat close to 0.5. Within patients, keeping preictal blocks intact and in time order cost the same spectral baseline 0.24 AUC on average compared with random allocation. These are different generalization tasks, and we do not merge them into one effect size. Taken together, they mean that a reported AUC is hard to interpret unless one knows across which boundary it was measured.

The cold-start result is also useful as a reference point. Patient-specific, calibrated or adaptive models address a different use case in which target data are available [13, 14]. Tested on the released partitions, such models can show directly how much target information adds to a model that has never seen the patient. The provenance map showed why the original index allocation did not separate episodes. Ordering windows and cutting the array by count let windows from the same episode fall into different partitions, and in all ten eligible cases raw samples also overlapped across a boundary. The chronological control removed those crossings and random allocation still scored higher. Randomization also changes which periods of a recording end up in training and in testing, so the difference cannot be attributed to overlapping samples alone. Event membership, time order and sample overlap are better treated as three separate checks.

We used PSD+LDA for the chronological control because its features and fitting rule were fixed and it needed no checkpoint selection. The control therefore says nothing about how EEGNet, TCN, 1D-CNN or EEG-Conformer would rank under complete-block temporal evaluation. Keeping every partition usable also meant a different cohort and variable window fractions. Results from such a control need to be reported with their eligibility rules and actual partition sizes, so readers can separate the effect of the allocation rule from a change in the cohort.

A false-positive cap of 1.67% on validation data did not guarantee the same proportion in new subjects, and window sensitivity at those thresholds was low, something an AUC summary would not reveal. This is evidence about how a fixed operating policy transfers under the sampled-window protocol, not an estimate of the false-alarm burden a patient would experience. A continuous warning evaluation would need causal preprocessing, a decision schedule, rules for alarm persistence or merging, a refractory period and measured time in warning [5, 15].

Our findings agree with earlier work on leakage and patient generalization [6, 7]. What this study adds is a window-to-recording audit, a temporal control matched on class counts and a validation-only threshold transfer analysis, all run on the same five pipelines and subject partitions. Adaptation studies address the case in which target data can inform the system [13, 14], and domain-generalization approaches aim to transfer seizure-prediction models without target-patient adaptation [27]. Differences in cohort, preprocessing, label intervals and target access rule out a direct ranking against the present results.

Some practical steps follow. An allocation manifest should identify the subject, recording, seizure block and sample interval of each window, so that shared samples and event membership can be checked before a classifier is fitted. The allocation rule should follow from the intended use: testing a person with no prior data requires subject separation, whereas testing later episodes within a person requires temporal separation. A threshold chosen on validation data should be carried unchanged into testing, with both the observed false-positive proportion and the sensitivity reported. A continuous warning study would also need measured monitoring time and a specified alarm policy.

The study has clear limits. The primary dataset is small and pediatric, and under the window construction and exclusion rules usable preictal windows represented only 69 of 182 annotated seizures. Class-dependent sampling changes the distribution of windows and does not reproduce continuous surveillance. We did not assess performance by sex or age. Filtering was noncausal, and the reconstructed timing relies on public metadata and the released fallback rules. Repeated partitions share subjects and fitted training sets, which limits population inference from their variability. The Siena check covers one baseline only. The temporal and threshold controls were added after the original analyses and are exploratory. A future evaluation should lock the complete signal and alarm pipeline before testing it on continuous external recordings, with an appropriate temporal null analysis [26].

## Conclusion

The same five EEG pipelines scored close to the 0.5 reference on unseen subjects, much higher under random window allocation than under chronological seizure blocks, and drifted from their intended operating points when validation thresholds were transferred to new people. Evaluation design deserves the same care in reporting as model design: which subjects, seizure episodes and samples were shared between training and testing, and where the threshold came from. The released partitions, window manifests and predictions allow new models to be compared with these results on the same terms.

## Supporting information

Reproducibility archive (code, manifests and predictions)

## Data Availability

This study used only publicly available, de-identified EEG data: the CHB-MIT Scalp EEG Database v1.0.0 (https://physionet.org/content/chbmit/1.0.0/) and the Siena Scalp EEG Database v1.0.0 (https://physionet.org/content/siena-scalp-eeg/1.0.0/), both on PhysioNet. No new clinical data were generated. Raw EDF files and full derived EEG arrays are not redistributed but can be obtained from PhysioNet and regenerated with the released preprocessing code. Preprocessing, model and analysis code is available at https://github.com/TsikonSong/EEG-Seizure-Prediction. Partition manifests, window maps, source tables and derived prediction arrays are provided in the reproducibility archive supplied as supplementary material.

https://physionet.org/content/siena-scalp-eeg/1.0.0/

https://github.com/TsikonSong/EEG-Seizure-Prediction

## Data availability

CHB-MIT version 1.0.0 and Siena version 1.0.0 are available from PhysioNet at https://physionet.org/content/chbmit/1.0.0/ and https://physionet.org/content/siena-scalp-eeg/1.0.0/, respectively. The study created no new clinical dataset. Source tables, partition manifests and derived score arrays accompany the manuscript. Raw EDFs and full derived EEG arrays are not redistributed; they can be obtained or regenerated from the public datasets using the released preprocessing code.

## Code availability

The original preprocessing, model and analysis code is available at https://github.com/TsikonSong/EEG-Seizure-Prediction, with the original benchmark audited against commit 845e973. The analyses added after the original benchmark, their recorded rules, runtime versions and derived results are supplied separately in the accompanying reproducibility archive and are not part of that commit. The tables and LaTeX figure sources allow the numerical summaries to be verified without the full EEG arrays.

## Ethics

This secondary analysis used the publicly available CHB-MIT and Siena databases. CHB-MIT protected health information was replaced with surrogate information [16]. Under Section 6.2 of the Auckland University of Technology Ethics Committee guidelines for existing published documents or datasets, no additional institutional ethics approval was required [28]. No new participants were recruited, no identifying information was sought and records were not linked between the two databases. No additional participant consent was required for this secondary analysis.

## Author contributions

Z.S.: Conceptualization, Methodology, Software, Formal analysis, Investigation, Data curation, Visualization, Writing – original draft, Writing – review and editing. M.P.: Supervision, Conceptualization, Methodology, Writing – review and editing. D.H.: Supervision, Conceptualization, Writing – review and editing. All authors approved the final manuscript.

## Supplementary material

The Supplementary Information contains cohort and allocation audits, numerical controls, threshold diagnostics and a reporting checklist. The reproducibility archive contains analysis code, source data and LaTeX sources for the manuscript and figures.

## Use of AI-assisted tools

Anthropic’s Claude was used for language editing and consistency checks. The authors take full responsibility for the content.

## Acknowledgments

We thank the CHB-MIT and Siena database teams and PhysioNet for making the recordings available for research.

