## Supplementary material for "Same pipelines, different answers: how evaluation design shapes EEG seizure prediction": Reproducibility archive (code, manifests and predictions): ANALYSIS_PROTOCOL.md

# Revision analyses fixed before their outcome summaries

Recorded 13 September 2026. These are exploratory additions during manuscript revision, not preregistered analyses. Original training and held-out score arrays are retained.

1. Frozen-model threshold transfer: use all 20 released subject-group partitions for all five models. Recover validation scores from saved deep-model weights, and reconstruct PSD+LDA from the original cached features. Check recovered test scores against archived predictions. Select the smallest representable threshold for which validation interictal FPR is <= 1/60 (>= decision rule, conservative ties). Report actual test FPR, sensitivity and partition distributions; never tune the rule on the test set. The cap is an illustrative window-level operating constraint, not a clinical standard.
2. Timing provenance: rerun candidate window indexing from raw EDF headers and original seizure annotations using released preprocessing logic. Accept provenance only when counts and labels match the saved data; verify representative stored waveforms against filtered source EEG. Retain all event memberships where preictal intervals overlap. Do not identify event counts from contiguous label runs.
3. Temporal sensitivity control: use PSD+LDA as the fixed transparent baseline (no hyperparameter search or checkpoint selection required). Compare (a) original file-index 60/20/20 allocation, (b) actual reconstructed-time 60/20/20 allocation and (c) chronological event-blocked allocation. Merge seizure episodes linked by shared preictal windows or overlapping preictal time intervals into indivisible blocks. Choose two time boundaries between blocks closest to the 60%/80% cumulative window targets, subject to both classes and at least one complete preictal block in every partition. Purge windows within 10 seconds of either boundary; report actual allocation fractions and the retained cohort. Match an additional 20 stratified random window splits to the event-blocked train/validation/test class counts in each eligible case, using the released seed list. Report per-case outcomes and descriptive paired differences. Eligibility and boundary selection use timing/labels, never model scores. This control isolates allocation behavior for one baseline and does not constitute a five-model prospective experiment.
4. Report all eligible cases, failures, effective cohort sizes and deviations from this protocol. No selective retention based on performance. Do not interpret repeated splits as independent patients, and do not convert sampled decisions into measured alarm rates.
