## Supplementary material for "Same pipelines, different answers: how evaluation design shapes EEG seizure prediction": Reproducibility archive (code, manifests and predictions): Figure_1.tex

\documentclass[tikz,border=4pt]{standalone}
\usepackage[T1]{fontenc}\usepackage{newtxtext,newtxmath}\usepackage{pgfplots}
\pgfplotsset{compat=1.18}\usepgfplotslibrary{groupplots}\usetikzlibrary{arrows.meta,positioning,calc}
\definecolor{modelblue}{HTML}{0072B2}\definecolor{modelorange}{HTML}{E69F00}
\definecolor{modelgreen}{HTML}{009E73}\definecolor{modelpurple}{HTML}{CC79A7}
\definecolor{modelred}{HTML}{D55E00}\definecolor{softgray}{HTML}{F2F3F5}
\begin{document}\input{figures/figure1_design}\end{document}
