## Supplementary material for "Same pipelines, different answers: how evaluation design shapes EEG seizure prediction": Reproducibility archive (code, manifests and predictions): README.md

# Figure data

These compact tables contain the exact observations plotted by the
LaTeX/PGFPlots figures. No points were simulated or reconstructed.

- tier3_per_seed.csv: 20 strict subject-grouped CHB-MIT partitions for each model.
- tier1_summary.csv: direct-overlap software stress-test results, including the EEGNet replication count.
- tier2_paired.csv: the earlier paired index-cut and random within-case diagnostic for ten cases. The index cut did not fully separate event blocks; this is distinct from the complete-block control.
- event_blocked_cases.csv: complete-block chronological and matched random control summaries for eleven cases, including eligible window counts.
- auc_means.csv: pooled and equally weighted subject-group AUC summaries used in the main comparison figure.
- validation_threshold_means.csv: held-out false-positive proportions and sensitivities summarised across the twenty validation-only threshold fits per model.
- validation_threshold_points.csv: the individual held-out operating points for those fits; thresholds were selected from validation scores and then fixed for testing.
- subject_balanced_per_group.csv: subject-group AUCs averaged over all strict test appearances, with each of 22 subject groups contributing equally.
- strict_low_far_per_seed.csv: post hoc, test-selected low-decision operating points from the strict subject-grouped prediction arrays. These are descriptive upper-bound diagnostics, not deployable validation-selected thresholds.
- strict_partition_manifest.csv: the exact training, validation and test case identifiers for all 20 strict seeds; chb01 and chb21 always move together.
- siena_strict_psd.csv: strict PSD+LDA CHB-MIT test AUCs and direct Siena transfer results, paired by seed.

The tables are deliberately small, auditable plotting inputs. The strict
low-decision table can be regenerated from the checked-in held-out prediction
archives with `work_J_far_constrained_sensitivity.py`. The Siena table can be
regenerated from the public CHB-MIT and Siena datasets with
`work_L_siena_external_psd_lda.py`. Neither path requires or modifies raw data
inside this repository.
