## Supplementary material for "Same pipelines, different answers: how evaluation design shapes EEG seizure prediction": Reproducibility archive (code, manifests and predictions): figure1_design.tex

% Figure 1: schematic of the three evaluation designs. Drawn with TikZ; no data are plotted.
\begin{tikzpicture}[font=\small,
panel/.style={draw=black!25,rounded corners=2pt,fill=softgray,minimum width=5.05cm,minimum height=4.55cm},
ttl/.style={font=\small\bfseries,anchor=north west},
sub/.style={font=\footnotesize,anchor=north west,text=black!75},
note/.style={font=\scriptsize,text=black!70,align=center},
sq/.style={minimum size=.34cm,inner sep=0pt,draw=white,line width=.6pt},
tr/.style={fill=modelblue!70},va/.style={fill=modelorange!80},te/.style={fill=modelred!75},
flow/.style={-{Latex[length=1.6mm]},semithick,draw=black!60}]
% ---------------- panel frames ----------------
\node[panel,anchor=north west] (PA) at (0,0) {};
\node[panel,anchor=north west] (PB) at (5.25,0) {};
\node[panel,anchor=north west] (PC) at (10.50,0) {};
% ---------------- (A) unseen subject groups ----------------
\node[ttl] at (0.12,-0.10) {(A) Unseen subject groups};
\node[sub] at (0.12,-0.52) {22 groups, 20 random partitions};
\foreach \i [count=\k from 0] in {tr,tr,te,tr,va,tr,tr,tr,te,tr,tr}{
  \node[sq,\i] at ({0.62+0.39*\k},-1.30) {};}
\foreach \i [count=\k from 0] in {va,tr,tr,te,tr,va,tr,te,tr,va,tr}{
  \node[sq,\i] at ({0.62+0.39*\k},-1.71) {};}
\draw[black,line width=.9pt] (0.62-0.17,-1.30-0.17) rectangle (0.62+0.17,-1.30+0.17);
\node[note,anchor=north west,align=left] at (0.12,-2.05) {\tikz\fill[modelblue!70] (0,0) rectangle (.2,.2); 14 train\quad
\tikz\fill[modelorange!80] (0,0) rectangle (.2,.2); 4 validation\quad
\tikz\fill[modelred!75] (0,0) rectangle (.2,.2); 4 test};
\node[note,anchor=north west,align=left] at (0.12,-2.55) {Outlined square: chb01 and chb21,\\one person, kept together};
\node[note,anchor=north west,align=left,text=black] at (0.12,-3.40) {Five pipelines. No data from the\\test subjects at any stage.};
% ---------------- (B) one patient over time ----------------
\node[ttl] at (5.37,-0.10) {(B) Within one patient};
\node[sub] at (5.37,-0.52) {PSD+LDA, 11 cases};
% preictal blocks (shaded) and seizure onsets; window k sits at x = 6.30 + 0.15k
\foreach \a/\b in {2/4,13/15,20/22}{
  \fill[black!12] ({6.30+0.15*\a-0.03},-1.05) rectangle ({6.30+0.15*\b+0.15},-2.62);
  \draw[modelred,line width=1.1pt] ({6.30+0.15*\b+0.21},-1.05)--({6.30+0.15*\b+0.21},-2.62);}
\node[font=\scriptsize,anchor=east] at (6.24,-1.42) {random};
\node[font=\scriptsize,anchor=east] at (6.24,-2.22) {blocks};
% random allocation row: 24 windows coloured at random
\foreach \c [count=\k from 0] in {tr,va,tr,te,tr,tr,va,te,tr,tr,te,tr,va,tr,tr,te,tr,va,tr,tr,te,tr,va,tr}{
  \fill[\c] ({6.30+0.15*\k},-1.30) rectangle ({6.30+0.15*\k+0.12},-1.54);}
% chronological row: earlier train, then validation, then later test; whole blocks stay together
\foreach \k in {0,...,10}{\fill[tr] ({6.30+0.15*\k},-2.10) rectangle ({6.30+0.15*\k+0.12},-2.34);}
\foreach \k in {12,...,16}{\fill[va] ({6.30+0.15*\k},-2.10) rectangle ({6.30+0.15*\k+0.12},-2.34);}
\foreach \k in {18,...,23}{\fill[te] ({6.30+0.15*\k},-2.10) rectangle ({6.30+0.15*\k+0.12},-2.34);}
\draw[flow] (6.30,-2.85)--(9.90,-2.85) node[right,font=\scriptsize,inner sep=1pt] {time};
\node[note,anchor=north west,align=left] at (5.37,-3.05) {Grey: preictal block, never split.\\Red line: seizure onset.};
\node[note,anchor=north west,align=left,text=black] at (5.37,-3.62) {Same windows and class counts;\\only the allocation rule changes.};
% ---------------- (C) threshold transfer ----------------
\node[ttl] at (10.62,-0.10) {(C) Threshold transfer};
\node[sub] at (10.62,-0.52) {Five pipelines, frozen fits};
\node[font=\scriptsize,anchor=west] at (10.62,-1.12) {validation interictal scores};
\draw[black!45] (10.75,-1.50)--(15.30,-1.50);
\foreach \x in {10.85,11.05,11.20,11.32,11.48,11.60,11.75,11.88,12.02,12.20,12.35,12.55,12.72,12.95,13.20,13.55,14.70}{
  \fill[modelorange] (\x,-1.50) circle (1.6pt);}
\node[font=\scriptsize,anchor=west] at (10.62,-1.92) {test interictal scores, new subjects};
\draw[black!45] (10.75,-2.40)--(15.30,-2.40);
\foreach \x in {10.90,11.10,11.30,11.52,11.70,11.95,12.15,12.40,12.62,12.90,13.25,13.70,14.20,14.55,14.95}{
  \fill[modelred] (\x,-2.40) circle (1.6pt);}
\draw[black,dashed,semithick] (14.40,-1.30)--(14.40,-2.62);
\node[font=\scriptsize,anchor=south] at (14.40,-1.32) {fixed $t$};
\node[note,anchor=north west,align=left] at (10.62,-3.05) {Dashed line: decision threshold $t$.};
\node[note,anchor=north west,align=left,text=black] at (10.62,-3.40) {$t$ lets $\leq1/60$ validation windows\\through, then is reused unchanged\\on the test subjects.};
\end{tikzpicture}
