## Supplementary material for "Same pipelines, different answers: how evaluation design shapes EEG seizure prediction": Reproducibility archive (code, manifests and predictions): figure3_temporal.tex

\begin{tikzpicture}\begin{axis}[width=15.3cm,height=6.3cm,xmin=.5,xmax=11.5,ymin=.2,ymax=1,
ylabel={AUC},xtick={1,...,11},xticklabels={chb01,chb04,chb05,chb06,chb08,chb09,chb10,chb13,chb14,chb15,chb22},tick label style={font=\small},label style={font=\small},
ymajorgrids,grid style={gray!18},axis line style={black!55},legend style={at={(.5,1.04)},anchor=south,draw=none,font=\small,legend columns=2}]
\addplot[black!50,dashed,forget plot] coordinates {(.5,.5)(11.5,.5)};
\addplot[black!35,forget plot] coordinates {(0.9,0.7244038155802861)(1.1,0.8479878775834656)};
\addplot[black!35,forget plot] coordinates {(1.9,0.3367745378953873)(2.1,0.9054555382324848)};
\addplot[black!35,forget plot] coordinates {(2.9,0.7197203852788958)(3.1,0.8375458237492811)};
\addplot[black!35,forget plot] coordinates {(3.9,0.5772804054054055)(4.1,0.7564294763513514)};
\addplot[black!35,forget plot] coordinates {(4.9,0.6097972972972974)(5.1,0.7177111486486486)};
\addplot[black!35,forget plot] coordinates {(5.9,0.5510654885654885)(6.1,0.8629781704781706)};
\addplot[black!35,forget plot] coordinates {(6.9,0.7663570691434468)(7.1,0.8591744066047473)};
\addplot[black!35,forget plot] coordinates {(7.9,0.2857142857142857)(8.1,0.8863095238095239)};
\addplot[black!35,forget plot] coordinates {(8.9,0.5853040540540541)(9.1,0.6096202572765075)};
\addplot[black!35,forget plot] coordinates {(9.9,0.3784786641929499)(10.1,0.7944495980210267)};
\addplot[black!35,forget plot] coordinates {(10.9,0.7558479532163742)(11.1,0.8885818713450295)};
\addplot[only marks,mark=*,mark size=2.5pt,modelblue] table[x expr=\thisrow{case_index}-.10,y=event_blocked_auc,col sep=comma]{figure_data/event_blocked_cases.csv};\addlegendentry{Chronological blocks}
\addplot[only marks,mark=triangle*,mark size=2.7pt,modelorange] table[x expr=\thisrow{case_index}+.10,y=random_mean_auc,col sep=comma]{figure_data/event_blocked_cases.csv};\addlegendentry{Matched random windows}
\end{axis}\end{tikzpicture}
