## Supplementary material for "Same pipelines, different answers: how evaluation design shapes EEG seizure prediction": Reproducibility archive (code, manifests and predictions): figure4_threshold.tex

\begin{tikzpicture}\begin{groupplot}[group style={group size=2 by 1,horizontal sep=1.75cm},width=7.2cm,height=6.0cm,
xmin=.65,xmax=5.35,xtick={1,2,3,4,5},xticklabels={PSD+LDA,EEGNet,TCN,1D-CNN,Conformer},
xticklabel style={rotate=32,anchor=east,font=\scriptsize},tick label style={font=\scriptsize},yticklabel style={/pgf/number format/fixed,/pgf/number format/precision=2},
label style={font=\small},title style={font=\small\bfseries},ymajorgrids,grid style={gray!18},axis line style={black!55}]
\nextgroupplot[title={(A) False-positive proportion},ylabel={Test interictal FPR},ymin=-.006,ymax=0.165]
\addplot[black!60,dashed] coordinates {(.65,.0166666667)(5.35,.0166666667)};
\addplot+[only marks,mark=*,mark size=1.5pt,modelblue,mark options={solid,fill=modelblue,draw=modelblue},restrict expr to domain={\thisrow{model_index}}{1:1}] table[x=x,y=test_fpr,col sep=comma] {figure_data/validation_threshold_points.csv};
\addplot+[only marks,mark=*,mark size=1.5pt,modelorange,mark options={solid,fill=modelorange,draw=modelorange},restrict expr to domain={\thisrow{model_index}}{2:2}] table[x=x,y=test_fpr,col sep=comma] {figure_data/validation_threshold_points.csv};
\addplot+[only marks,mark=*,mark size=1.5pt,modelgreen,mark options={solid,fill=modelgreen,draw=modelgreen},restrict expr to domain={\thisrow{model_index}}{3:3}] table[x=x,y=test_fpr,col sep=comma] {figure_data/validation_threshold_points.csv};
\addplot+[only marks,mark=*,mark size=1.5pt,modelpurple,mark options={solid,fill=modelpurple,draw=modelpurple},restrict expr to domain={\thisrow{model_index}}{4:4}] table[x=x,y=test_fpr,col sep=comma] {figure_data/validation_threshold_points.csv};
\addplot+[only marks,mark=*,mark size=1.5pt,modelred,mark options={solid,fill=modelred,draw=modelred},restrict expr to domain={\thisrow{model_index}}{5:5}] table[x=x,y=test_fpr,col sep=comma] {figure_data/validation_threshold_points.csv};
\addplot[only marks,mark=square*,mark size=2.2pt,black] table[x=model_index,y=test_fpr_mean,col sep=comma]{figure_data/validation_threshold_means.csv};
\nextgroupplot[title={(B) Window sensitivity},ylabel={Test preictal sensitivity},ymin=-.006,ymax=0.28]
\addplot+[only marks,mark=*,mark size=1.5pt,modelblue,mark options={solid,fill=modelblue,draw=modelblue},restrict expr to domain={\thisrow{model_index}}{1:1}] table[x=x,y=test_sensitivity,col sep=comma] {figure_data/validation_threshold_points.csv};
\addplot+[only marks,mark=*,mark size=1.5pt,modelorange,mark options={solid,fill=modelorange,draw=modelorange},restrict expr to domain={\thisrow{model_index}}{2:2}] table[x=x,y=test_sensitivity,col sep=comma] {figure_data/validation_threshold_points.csv};
\addplot+[only marks,mark=*,mark size=1.5pt,modelgreen,mark options={solid,fill=modelgreen,draw=modelgreen},restrict expr to domain={\thisrow{model_index}}{3:3}] table[x=x,y=test_sensitivity,col sep=comma] {figure_data/validation_threshold_points.csv};
\addplot+[only marks,mark=*,mark size=1.5pt,modelpurple,mark options={solid,fill=modelpurple,draw=modelpurple},restrict expr to domain={\thisrow{model_index}}{4:4}] table[x=x,y=test_sensitivity,col sep=comma] {figure_data/validation_threshold_points.csv};
\addplot+[only marks,mark=*,mark size=1.5pt,modelred,mark options={solid,fill=modelred,draw=modelred},restrict expr to domain={\thisrow{model_index}}{5:5}] table[x=x,y=test_sensitivity,col sep=comma] {figure_data/validation_threshold_points.csv};
\addplot[only marks,mark=square*,mark size=2.2pt,black] table[x=model_index,y=test_sensitivity_mean,col sep=comma]{figure_data/validation_threshold_means.csv};
\end{groupplot}\end{tikzpicture}
