## Supplementary material for "Same pipelines, different answers: how evaluation design shapes EEG seizure prediction": Reproducibility archive (code, manifests and predictions): preamble.tex

\usepackage[T1]{fontenc}
\usepackage[utf8]{inputenc}
\usepackage[a4paper,margin=2.35cm]{geometry}
\usepackage{newtxtext,newtxmath}
\usepackage{amsmath}
\usepackage{booktabs}
\usepackage{threeparttable}
\usepackage{tabularx}
\usepackage{graphicx}
\usepackage{placeins}
\usepackage{enumitem}
\usepackage{tikz}
\usepackage{microtype}
\usepackage{xcolor}
\usepackage{pgfplots}
\usepackage[
  backend=biber,
  style=numeric-comp,
  sorting=none,
  maxbibnames=6,
  minbibnames=6,
  giveninits=true,
  doi=true,
  url=true,
  eprint=true,
  isbn=false,
  natbib=true
]{biblatex}
\addbibresource{references.bib}
% Vancouver-style bibliography: surname and initials; journal year;volume:pages.
\DeclareNameAlias{author}{family-given}
\renewcommand*{\revsdnamepunct}{}
\renewcommand*{\bibinitperiod}{}
\renewcommand*{\bibinitdelim}{}
\DeclareDelimFormat{multinamedelim}{\addcomma\space}
\DeclareDelimFormat{finalnamedelim}{\addcomma\space}
\DeclareFieldFormat[article,misc,online,thesis]{title}{#1}
\DeclareFieldFormat{url}{\url{#1}}
\DeclareFieldFormat{journaltitle}{#1}
\DeclareFieldFormat{pages}{#1}
\DeclareFieldFormat{labelnumberwidth}{#1\addperiod}
\DeclareBibliographyDriver{article}{%
  \printnames{author}\setunit{\addperiod\space}%
  \printfield{title}\setunit{\addperiod\space}%
  \printfield{journaltitle}\setunit{\addspace}\printfield{year}%
  \iffieldundef{volume}{}{\setunit{\addsemicolon}\printfield{volume}}%
  \iffieldundef{number}{}{\printtext[parens]{\printfield{number}}}%
  \iffieldundef{pages}{}{\setunit{\addcolon}\printfield{pages}}%
  \newunit\newblock\printfield{doi}\newunit\printfield{note}\usebibmacro{finentry}}
\DeclareBibliographyDriver{misc}{%
  \printnames{author}\setunit{\addperiod\space}\printfield{title}%
  \setunit{\addperiod\space}\printfield{howpublished}%
  \setunit{\addsemicolon\space}\printfield{year}%
  \newunit\newblock\printfield{note}%
  \newunit\newblock\printfield{doi}\usebibmacro{finentry}}
\DeclareBibliographyDriver{online}{%
  \printnames{author}\setunit{\addperiod\space}\printfield{title}%
  \setunit{\addspace}\printtext{[Internet]}%
  \newunit\printtext{[cited \printfield{urlyear}\space\mkbibmonth{\thefield{urlmonth}}\space\printfield{urlday}]}%
  \newunit\printtext{Available from: }\printfield{url}%
  \usebibmacro{finentry}}
\DeclareBibliographyDriver{thesis}{%
  \printnames{author}\setunit{\addperiod\space}\printfield{title}%
  \setunit{\addspace}\printtext{[PhD thesis]}%
  \newunit\printlist{institution}\setunit{\addsemicolon\space}\printfield{year}%
  \newunit\printfield{url}\usebibmacro{finentry}}
\usepackage[switch]{lineno}
\usepackage{caption}
\usepackage{url}
\usepackage[hidelinks]{hyperref}
\Urlmuskip=0mu plus 2mu\relax

\pgfplotsset{compat=1.18}
\usepgfplotslibrary{groupplots}
\usetikzlibrary{arrows.meta,positioning,fit,backgrounds,calc}
\definecolor{modelblue}{HTML}{0072B2}
\definecolor{modelorange}{HTML}{E69F00}
\definecolor{modelgreen}{HTML}{009E73}
\definecolor{modelpurple}{HTML}{CC79A7}
\definecolor{modelred}{HTML}{D55E00}
\definecolor{softgray}{HTML}{F2F3F5}
\captionsetup{font=small,labelfont=bf}
\modulolinenumbers[5]
\raggedbottom
% Unnumbered headings; figure and table numbering is retained.
\setcounter{secnumdepth}{0}
