## Supplementary material for "Same pipelines, different answers: how evaluation design shapes EEG seizure prediction": Reproducibility archive (code, manifests and predictions): README.md

# Manuscript source

LaTeX source of the preprint "Same pipelines, different answers: how evaluation
design shapes EEG seizure prediction" (Song, Pedersen and Henare), with its
supplementary information, bibliography, tables and figure sources.

Compile with a recent TeX distribution including biblatex, Biber and PGFPlots:

```sh
latexmk -pdf main.tex
latexmk -pdf supplement.tex
latexmk -pdf Figure_1.tex Figure_2.tex Figure_3.tex Figure_4.tex
```

The supplied main.bbl and supplement.bbl contain the resolved bibliography.
Figure data are documented in figure_data/README.md. Every figure is drawn with
TikZ/PGFPlots from the source tables in figure_data/; no image generator was used.
