## Supplementary material for "Same pipelines, different answers: how evaluation design shapes EEG seizure prediction": Reproducibility archive (code, manifests and predictions): references.bib

@online{autec2026,
  author  = {{Auckland University of Technology}},
  title   = {Exemptions to activities requiring {AUT} Ethics Committee approval (6)},
  url     = {https://www.aut.ac.nz/research/researchethics/autec-guidelines-and-procedures/exemptions-to-activities-requiring-aut-ethics-committee-approval-6},
  urldate = {2026-10-05},
  note    = {Section 6.2: existing published documents or datasets}
}

@article{andrade2024,
  author  = {Andrade, In{\^e}s and Teixeira, C{\'e}sar and Pinto, Mauro},
  title   = {On the performance of seizure prediction machine learning methods across different databases: The sample and alarm-based perspectives},
  journal = {Front. Neurosci.},
  year    = {2024},
  volume  = {18},
  pages   = {1417748},
  doi     = {10.3389/fnins.2024.1417748}
}

@article{collins2024tripodai,
  author  = {Collins, Gary S. and Moons, Karel G. M. and Dhiman, Paula and Riley, Richard D. and Beam, Andrew L. and Van Calster, Ben and Ghassemi, Marzyeh and Liu, Xiaoxuan and Reitsma, Johannes B. and van Smeden, Maarten and Boulesteix, Anne-Laure and others},
  title   = {{TRIPOD+AI} statement: updated guidance for reporting clinical prediction models that use regression or machine learning methods},
  journal = {BMJ},
  year    = {2024},
  volume  = {385},
  pages   = {e078378},
  doi     = {10.1136/bmj-2023-078378}
}

@article{moons2025probastai,
  author  = {Moons, Karel G. M. and Damen, Johanna A. A. and Kaul, Tabea and Hooft, Lotty and Andaur Navarro, Constanza and Dhiman, Paula and Beam, Andrew L. and Van Calster, Ben and Celi, Leo Anthony and Denaxas, Spiros and others},
  title   = {{PROBAST+AI}: An updated quality, risk of bias, and applicability assessment tool for prediction models using regression or artificial intelligence methods},
  journal = {BMJ},
  year    = {2025},
  volume  = {388},
  pages   = {e082505},
  doi     = {10.1136/bmj-2024-082505}
}

@misc{who2024epilepsy,
  author  = {{World Health Organization}},
  title   = {Epilepsy},
  date    = {2024-02-07},
  url     = {https://www.who.int/news-room/fact-sheets/detail/epilepsy},
  urldate = {2026-08-17}
}

@article{andrzejak2026,
  author  = {Andrzejak, Ralph G. and Bre{\v{s}}ar, Martin and Richardson, Mark P. and Viana, Pedro F. and Rosch, Richard},
  title   = {Are seizure forecasts and cycles better than chance? What chance?},
  journal = {Epilepsia},
  year    = {2026},
  volume  = {67},
  number  = {6},
  pages   = {2966--2978},
  doi     = {10.1002/epi.70158}
}

@article{bai2018,
  author  = {Bai, Shaojie and Kolter, J. Zico and Koltun, Vladlen},
  title   = {An empirical evaluation of generic convolutional and recurrent networks for sequence modeling},
  journal = {arXiv},
  year    = {2018},
  eprint  = {1803.01271},
  doi     = {10.48550/arXiv.1803.01271}
}

@article{brookshire2024,
  author  = {Brookshire, Geoffrey and Kasper, Jake and Blauch, Nicholas M. and Wu, Yunan Charles and Glatt, Ryan and Merrill, David A. and Gerrol, Spencer and Yoder, Keith J. and Quirk, Colin and Lucero, Ch{\'e}},
  title   = {Data leakage in deep learning studies of translational {EEG}},
  journal = {Front. Neurosci.},
  year    = {2024},
  volume  = {18},
  pages   = {1373515},
  doi     = {10.3389/fnins.2024.1373515}
}

@article{carmo2024,
  author  = {Carmo, Ana Sofia and Abreu, Mariana and Baptista, Maria Fortuna and de Oliveira Carvalho, Miguel and Peralta, Ana Rita and Fred, Ana and Bentes, Carla and da Silva, Hugo Plácido},
  title   = {Automated algorithms for seizure forecast: A systematic review and meta-analysis},
  journal = {J. Neurol.},
  year    = {2024},
  volume  = {271},
  number  = {10},
  pages   = {6573--6587},
  doi     = {10.1007/s00415-024-12655-z}
}

@article{goldberger2000,
  author  = {Goldberger, Ary L. and Amaral, Luis A. N. and Glass, Leon and Hausdorff, Jeffrey M. and Ivanov, Plamen Ch. and Mark, Roger G. and Mietus, Joseph E. and Moody, George B. and Peng, Chung-Kang and Stanley, H. Eugene},
  title   = {{PhysioBank}, {PhysioToolkit}, and {PhysioNet}: Components of a new research resource for complex physiologic signals},
  journal = {Circulation},
  year    = {2000},
  volume  = {101},
  number  = {23},
  pages   = {e215--e220},
  doi     = {10.1161/01.CIR.101.23.e215}
}

@article{pollard2026physionet,
  author  = {Pollard, Tom and Moody, Benjamin E. and Lehman, Li-wei H. and Gow, Brian J. and Fernandes, Chrystinne and Xie, Chen and Johnson, Alistair and Mark, Roger G. and Heldt, Thomas},
  title   = {{PhysioNet} as a global platform for biomedical research},
  journal = {Nat. Health},
  year    = {2026},
  volume  = {1},
  number  = {8},
  pages   = {792--795},
  doi     = {10.1038/s44360-026-00096-z}
}

@misc{guttag2010,
  author       = {Guttag, John},
  title        = {{CHB-MIT} Scalp {EEG} Database},
  year         = {2010},
  howpublished = {PhysioNet, version 1.0.0},
  doi          = {10.13026/C2K01R},
  url          = {https://physionet.org/content/chbmit/1.0.0/}
}

@article{jemal2024,
  author  = {Jemal, Imen and Abou-Abbas, Lina and Henni, Karim and Mitiche, Amar and Mezghani, Nizar},
  title   = {Domain adaptation for {EEG}-based, cross-subject epileptic seizure prediction},
  journal = {Front. Neuroinform.},
  year    = {2024},
  volume  = {18},
  pages   = {1303380},
  doi     = {10.3389/fninf.2024.1303380}
}

@article{zhang2025cda,
  author  = {Zhang, Ziye and Liu, Aiping and Gao, Yikai and Qian, Ruobing and Chen, Xun},
  title   = {Cross-patient seizure prediction via continuous domain adaptation and similar sample replay},
  journal = {Cogn. Neurodyn.},
  year    = {2025},
  volume  = {19},
  number  = {1},
  pages   = {26},
  doi     = {10.1007/s11571-024-10216-8}
}

@article{karoly2026,
  author  = {Karoly, Philippa J. and Stirling, Rachel E. and Cook, Mark J. and Goldenholz, Daniel M. and Baud, Maxime O. and Rao, Vikram R. and Vieluf, Solveig and Brinkmann, Benjamin H.},
  title   = {Seizure forecasting: The long and winding road to clinical translation},
  journal = {Epilepsia},
  year    = {2026},
  doi     = {10.1002/epi.70394},
  note    = {Published online 20 July 2026}
}

@article{kwan2000,
  author  = {Kwan, Patrick and Brodie, Martin J.},
  title   = {Early identification of refractory epilepsy},
  journal = {N. Engl. J. Med.},
  year    = {2000},
  volume  = {342},
  number  = {5},
  pages   = {314--319},
  doi     = {10.1056/NEJM200002033420503}
}

@article{lawhern2018,
  author  = {Lawhern, Vernon J. and Solon, Amelia J. and Waytowich, Nicholas R. and Gordon, Stephen M. and Hung, Chou P. and Lance, Brent J.},
  title   = {{EEGNet}: A compact convolutional neural network for {EEG}-based brain--computer interfaces},
  journal = {J. Neural Eng.},
  year    = {2018},
  volume  = {15},
  number  = {5},
  pages   = {056013},
  doi     = {10.1088/1741-2552/aace8c}
}

@article{mao2023,
  author  = {Mao, Tingting and Li, Chang and Zhao, Yuchang and Song, Rencheng and Chen, Xun},
  title   = {Online test-time adaptation for patient-independent seizure prediction},
  journal = {IEEE Sens. J.},
  year    = {2023},
  volume  = {23},
  number  = {19},
  pages   = {23133--23144},
  doi     = {10.1109/JSEN.2023.3307223}
}

@article{mormann2007,
  author  = {Mormann, Florian and Andrzejak, Ralph G. and Elger, Christian E. and Lehnertz, Klaus},
  title   = {Seizure prediction: The long and winding road},
  journal = {Brain},
  year    = {2007},
  volume  = {130},
  number  = {2},
  pages   = {314--333},
  doi     = {10.1093/brain/awl241}
}

@article{schulze2010,
  author  = {Schulze-Bonhage, Andreas and Sales, Francisco and Wagner, Kathrin and Teotonio, Rute and Carius, Astrid and Schelle, Annette and Ihle, Matthias},
  title   = {Views of patients with epilepsy on seizure prediction devices},
  journal = {Epilepsy Behav.},
  year    = {2010},
  volume  = {18},
  number  = {4},
  pages   = {388--396},
  doi     = {10.1016/j.yebeh.2010.05.008}
}

@article{segal2023,
  author  = {Segal, Galya and Keidar, Noam and Lotan, Roy Maor and Romano, Yaniv and Herskovitz, Moshe and Yaniv, Yael},
  title   = {Utilizing risk-controlling prediction calibration to reduce false alarm rates in epileptic seizure prediction},
  journal = {Front. Neurosci.},
  year    = {2023},
  volume  = {17},
  pages   = {1184990},
  doi     = {10.3389/fnins.2023.1184990}
}

@article{shafiezadeh2023,
  author  = {Shafiezadeh, Sina and Duma, Gian Marco and Mento, Giovanni and Danieli, Alberto and Antoniazzi, Lisa and Del Popolo Cristaldi, Fiorella and Bonanni, Paolo and Testolin, Alberto},
  title   = {Methodological issues in evaluating machine learning models for {EEG} seizure prediction: Good cross-validation accuracy does not guarantee generalization to new patients},
  journal = {Appl. Sci.},
  year    = {2023},
  volume  = {13},
  number  = {7},
  pages   = {4262},
  doi     = {10.3390/app13074262}
}

@article{shafiezadeh2024calibration,
  author  = {Shafiezadeh, Sina and Duma, Gian Marco and Mento, Giovanni and Danieli, Alberto and Antoniazzi, Lisa and Del Popolo Cristaldi, Fiorella and Bonanni, Paolo and Testolin, Alberto},
  title   = {Calibrating deep learning classifiers for patient-independent electroencephalogram seizure forecasting},
  journal = {Sensors},
  year    = {2024},
  volume  = {24},
  number  = {9},
  pages   = {2863},
  doi     = {10.3390/s24092863}
}

@article{shafiezadeh2024review,
  author  = {Shafiezadeh, Sina and Duma, Gian Marco and Pozza, Marco and Testolin, Alberto},
  title   = {A systematic review of cross-patient approaches for {EEG} epileptic seizure prediction},
  journal = {J. Neural Eng.},
  year    = {2024},
  volume  = {21},
  number  = {6},
  pages   = {061004},
  doi     = {10.1088/1741-2552/ad9682}
}

@phdthesis{shoeb2009,
  author = {Shoeb, Ali H.},
  title  = {Application of machine learning to epileptic seizure onset detection and treatment},
  school = {Harvard--MIT Division of Health Sciences and Technology},
  year   = {2009},
  url    = {https://hdl.handle.net/1721.1/54669}
}

@article{snyder2008,
  author  = {Snyder, David E. and Echauz, Javier and Grimes, David B. and Litt, Brian},
  title   = {The statistics of a practical seizure warning system},
  journal = {J. Neural Eng.},
  year    = {2008},
  volume  = {5},
  number  = {4},
  pages   = {392--401},
  doi     = {10.1088/1741-2560/5/4/004}
}

@misc{detti2020siena,
  author       = {Detti, Paolo},
  title        = {Siena Scalp {EEG} Database},
  year         = {2020},
  howpublished = {PhysioNet},
  note         = {Version 1.0.0},
  doi          = {10.13026/5d4a-j060},
  url          = {https://physionet.org/content/siena-scalp-eeg/1.0.0/}
}

@article{detti2020synchronization,
  author  = {Detti, Paolo and Vatti, Giampaolo and {Zabalo Manrique de Lara}, Garazi},
  title   = {{EEG} synchronization analysis for seizure prediction: A study on data of noninvasive recordings},
  journal = {Processes},
  year    = {2020},
  volume  = {8},
  number  = {7},
  pages   = {846},
  doi     = {10.3390/pr8070846}
}

@article{song2023conformer,
  author  = {Song, Yonghao and Zheng, Qingqing and Liu, Bingchuan and Gao, Xiaorong},
  title   = {{EEG Conformer}: Convolutional transformer for {EEG} decoding and visualization},
  journal = {IEEE Trans. Neural Syst. Rehabil. Eng.},
  year    = {2023},
  volume  = {31},
  pages   = {710--719},
  doi     = {10.1109/TNSRE.2022.3230250}
}

@article{song2026review,
  author  = {Song, Zikang and Arrowsmith, Kim and Henare, Dion and Pedersen, Mangor},
  title   = {Real-time {EEG}-based epileptic seizure prediction using artificial intelligence: A systematic review},
  journal = {Artif. Intell. Med.},
  year    = {2026},
  volume  = {180},
  pages   = {103476},
  doi     = {10.1016/j.artmed.2026.103476},
  note    = {Published online 26 June 2026}
}

@article{truong2018,
  author  = {Truong, Nhan Duy and Nguyen, Anh Duy and Kuhlmann, Levin and Bonyadi, Mohammad Reza and Yang, Jiawei and Ippolito, Stephen and Kavehei, Omid},
  title   = {Convolutional neural networks for seizure prediction using intracranial and scalp electroencephalogram},
  journal = {Neural Netw.},
  year    = {2018},
  volume  = {105},
  pages   = {104--111},
  doi     = {10.1016/j.neunet.2018.04.018}
}

@article{yang2026causal,
  author  = {Yang, Hongliu and M{\"u}ller, Jens and Kalousios, Sotirios and Leonhardt, Georg and D{\"u}mpelmann, Matthias and Schulze-Bonhage, Andreas and Tetzlaff, Ronald},
  title   = {Seizure forecasting with epilepsy cycles: On the causality of forecasting pipelines},
  journal = {Epilepsia},
  year    = {2026},
  volume  = {67},
  number  = {5},
  pages   = {2506--2517},
  doi     = {10.1002/epi.70110}
}

@article{zhao2026seifud,
  author  = {Zhao, Sha and Zhang, Caibo and Wang, Jiquan and Jiang, Haiteng and Li, Shijian and Li, Tao and Pan, Gang},
  title   = {{SeiFuD}: A novel deep learning framework leveraging domain generalization for cross-subject seizure prediction},
  journal = {Neurocomputing},
  year    = {2026},
  volume  = {666},
  pages   = {132227},
  doi     = {10.1016/j.neucom.2025.132227}
}
