## Supplementary material for "Same pipelines, different answers: how evaluation design shapes EEG seizure prediction": Reproducibility archive (code, manifests and predictions): results_macros.tex

\newcommand{\LowFprRange}{0.018--0.031}
\newcommand{\LowSenRange}{0.023--0.056}
\newcommand{\LowFprPercentRange}{1.81--3.13\%}
\newcommand{\LowSenPercentRange}{2.33--5.56\%}
\newcommand{\ExceedRange}{6--10}
