## Supplementary material for "Same pipelines, different answers: how evaluation design shapes EEG seizure prediction": Reproducibility archive (code, manifests and predictions): supplement.tex

\documentclass[11pt]{article}
\input{preamble}
\usepackage{float}
\renewcommand{\thetable}{S\arabic{table}}
\renewcommand{\theequation}{S\arabic{equation}}
\title{Supplementary Information\\[.4em]\large Same pipelines, different answers: how evaluation design shapes EEG seizure prediction}
\author{Zikang Song, Mangor Pedersen and Dion Henare\\\small Auckland University of Technology, Auckland, New Zealand}
\date{}
\hypersetup{pdftitle={Supplementary Information: Same pipelines, different answers: how evaluation design shapes EEG seizure prediction},pdfauthor={Zikang Song; Mangor Pedersen; Dion Henare}}
\begin{document}
\maketitle
\section{Cohort and provenance}
The common CHB-MIT cohort contains 23 case identifiers from 22 subject groups. Table~S1 separates annotation identities, retained windows and preictal blocks. The reconstructed map recovered all 17,424 saved windows. In chb17, seven additional candidates were rejected by the original flatline rule. Waveform checks found zero discrepancy for 126 selected windows from the other cases and a maximum absolute normalized-signal difference of $5.96\times10^{-8}$ across all 491 retained chb17 windows.
\input{tables/cohort}
\clearpage
\section{Subject allocation and chronological control}
Table~S2 reports how often each independent subject group appeared in testing. Table~S3 gives actual window counts for the chronological block control. Its eleven-case cohort comprises chb01, chb04, chb05, chb06, chb08, chb09, chb10, chb13, chb14, chb15 and chb22. Each includes at least three separable preictal blocks and both classes in every selected partition.
\input{tables/frequency}
\input{tables/blocked}
\clearpage
\section{Original index-allocation audit}
The original within-case scheme cut concatenated windows at 60\% and 80\% by index. Earlier versions of this analysis described this allocation as chronological, although it did not keep complete preictal episodes in one partition. Table~S4 identifies crossings from the reconstructed source map. Sorting the windows by reconstructed time did not change the PSD+LDA results in these ten cases.
\input{tables/boundaries}
\begin{table}[H]
\centering
\caption{Original five-pipeline matched random-versus-index comparison.}
\small
\begin{tabular}{lcccc}
\toprule
Pipeline & Index AUC & Random AUC & Difference & $p$\\
\midrule
PSD+LDA & $0.607\pm0.215$ & $0.841\pm0.097$ & 0.233 & 0.004\\
EEGNet & $0.558\pm0.158$ & $0.697\pm0.171$ & 0.139 & 0.105\\
TCN & $0.558\pm0.251$ & $0.871\pm0.115$ & 0.313 & 0.006\\
1D-CNN & $0.549\pm0.239$ & $0.922\pm0.063$ & 0.373 & 0.004\\
EEG-Conformer & $0.616\pm0.254$ & $0.885\pm0.117$ & 0.269 & 0.004\\
\bottomrule
\end{tabular}
\par\smallskip\footnotesize Values are mean $\pm$ SD across ten cases. Each random value first averages 20 stratified 60/20/20 window allocations. Differences and two-sided Wilcoxon signed-rank $p$ values use the case as the paired unit. The five-comparison Bonferroni threshold is 0.01. Both schemes permit related windows across partitions; this is not a comparison with complete-block validation.
\end{table}
\clearpage
\section{Threshold selection and numerical recovery}
Table~S6 reports the validation-only policy. Its rule was fixed before its outcome summary was inspected: allow at most $\lfloor N_0/60\rfloor$ positive decisions among $N_0$ validation interictal windows, with ties handled conservatively. Test labels were used only to calculate the held-out outcomes. The validation scores came from the saved checkpoints or reconstructed PSD+LDA fits; the test scores were the archived arrays from the original benchmark.
\input{tables/thresholds}
Recovered deep-model validation AUCs differed from the archived checkpoint records by less than $10^{-4}$. To check numerical sensitivity, we shifted each selected threshold upward and downward by that run's largest absolute recovered-versus-archived test-score discrepancy. Across pipelines and directions, the largest absolute change in a model's mean test FPR was 0.00042, and the largest change in mean window sensitivity was 0.00027. This is a perturbation check, not a bound on every possible numerical implementation difference.

The original cadence-adjusted quantity was
\begin{equation}
\mathrm{FPD}_{300}=\frac{\mathrm{FP}}{N_0\times300/3600}=12\,\mathrm{FPR}.
\end{equation}
It assigns a nominal five-minute interval to each interictal decision. It is an algebraic rescaling of sampled-window FPR and does not recover measured monitoring hours. Under the original validation Youden rule, its means were 4.994, 4.215, 5.337, 4.175 and 5.186 nominal decisions/h for PSD+LDA, EEGNet, TCN, 1D-CNN and EEG-Conformer, respectively. The main manuscript reports specificity instead.
\begin{table}[H]
\centering
\caption{Preserved test-selected low-decision diagnostic, separate from the validation policy.}
\small
\begin{tabular}{lcc}
\toprule
Pipeline & Nominal $\mathrm{FPD}_{300}$/h & Window sensitivity\\
\midrule
PSD+LDA & $0.195\pm0.003$ & $0.019\pm0.013$\\
EEGNet & $0.195\pm0.003$ & $0.023\pm0.017$\\
TCN & $0.195\pm0.003$ & $0.025\pm0.017$\\
1D-CNN & $0.195\pm0.003$ & $0.033\pm0.027$\\
EEG-Conformer & $0.195\pm0.003$ & $0.019\pm0.019$\\
\bottomrule
\end{tabular}
\par\smallskip\footnotesize Means $\pm$ SD across the 20 strict subject partitions. Test labels selected a threshold to maximize sensitivity subject to $\mathrm{FPD}_{300}\leq0.2$/h, equivalent to sampled-window FPR $\leq1/60$. These are post hoc score-separation results, not performance of a locked threshold transferred to unseen data. They are retained to trace the earlier analysis and are not the new primary operating-point result.
\end{table}
\clearpage
\section{External and deliberately contaminated controls}
The Siena analysis used PSD+LDA models fitted on the CHB-MIT training groups and thresholds from the corresponding CHB-MIT validation groups. The low-decision Siena threshold was part of the pre-existing external control; it was not selected on Siena test labels. This threshold was selected using CHB-MIT validation data, whereas the internal diagnostic in Table~S7 used CHB-MIT test data for threshold selection.
\begin{table}[H]
\centering
\caption{Existing CHB-MIT-to-Siena PSD+LDA portability control.}
\small
\begin{tabular}{lc}
\toprule
Metric & Mean $\pm$ SD\\
\midrule
CHB-MIT internal pooled AUC & $0.527\pm0.070$\\
Siena pooled AUC & $0.480\pm0.022$\\
Siena participant-macro AUC & $0.477\pm0.024$\\
Siena sensitivity at CHB-MIT validation Youden threshold & $0.195\pm0.145$\\
Siena nominal $\mathrm{FPD}_{300}$/h at that threshold & $2.600\pm1.794$\\
Siena sensitivity at CHB-MIT validation low-decision threshold & $0.013\pm0.020$\\
Siena nominal $\mathrm{FPD}_{300}$/h at that threshold & $0.238\pm0.298$\\
\bottomrule
\end{tabular}
\par\smallskip\footnotesize The 20 CHB-MIT fits were each applied to the same 13 Siena participants. The SD describes variation across these fits; it is not based on 20 independent external cohorts. All decision-rate quantities are nominal rescalings of sampled-window FPR.
\end{table}

The overlap stress test deliberately reconstructed a malformed pooled-window allocation. All preictal windows and a seed-specific 20\% of interictal windows were pooled. One stratified fold was assigned to testing and one to validation. Training concatenated the training complements of three other folds. Every validation and test sample therefore appeared three times in the training index; remaining samples appeared twice. This changes sample overlap, interictal selection and index multiplicity together.
\begin{table}[H]
\centering
\caption{Deliberate exact-sample-overlap stress test.}
\small
\begin{tabular}{lc}
\toprule
Pipeline & AUC\\
\midrule
PSD+LDA & 0.736\\
EEGNet & $0.964\pm0.010$\\
TCN & 1.000\\
1D-CNN & 1.000\\
EEG-Conformer & 0.9999\\
\bottomrule
\end{tabular}
\par\smallskip\footnotesize EEGNet is a five-seed mean $\pm$ SD; the other values are single runs. This software-integrity diagnostic is excluded from generalization inference. Its difference from subject holdout is not an isolated estimate of leakage bias.
\end{table}
\clearpage
\section{Information use and reproducibility}
Table~S10 records which information supports each step. The chronological and threshold analyses were added after the original benchmark and are exploratory. They did not change the original subject splits, model weights or archived held-out predictions.
\begin{table}[H]
\centering
\caption{Information-use audit.}
\small
\begin{tabularx}{\textwidth}{p{3.4cm}X}
\toprule
Step & Information used and restrictions\\
\midrule
Subject partitioning & Published case identities and the fixed seed list; chb01/chb21 remain together.\\
Signal normalization & Each individual window and channel; no statistics pooled across partitions. Zero-phase filtering remains retrospective.\\
Deep-model fitting & Training windows only; validation AUC selects the checkpoint and stopping epoch.\\
Original Youden threshold & Validation labels and scores, with the recorded 0.5 fallback when validation AUC is below 0.5.\\
Chronological boundaries & Reconstructed times, seizure membership and class availability; boundaries are selected without model scores.\\
Matched random control & The blocked split's retained windows and exact class counts; 20 fixed random seeds.\\
New low-FPR threshold & Validation interictal scores only; test data do not determine the threshold.\\
Post hoc low-decision diagnostic & Test scores and labels; retained solely as a test-selected diagnostic.\\
External PSD+LDA & CHB-MIT fitting and validation only; Siena is evaluated without target fitting or threshold selection.\\
Subject-balanced AUC & Archived test scores within group, averaged across repeated test appearances before equal weighting.\\
Overlap stress test & Evaluation samples deliberately reused during training; no deployment interpretation.\\
\bottomrule
\end{tabularx}
\end{table}

The later analyses used Python 3.12, PyTorch 2.5.1, MNE 1.13.2 and the package versions recorded in \texttt{runtime.json}. Deep-model score recovery used Apple MPS; the original checkpoints and held-out predictions were preserved. The reproducibility archive contains the scripts for the later analyses, allocation rules, window maps, per-case and per-partition results, validation predictions, original test predictions and PSD features. Paths can be set through environment variables. The original benchmark code is pinned separately to commit \texttt{845e973}.

The seed list is 42, 123, 456, 789, 1024, 2025, 3141, 4096, 5555, 6174, 7077, 8192, 9001, 9999, 11111, 12345, 13579, 14142, 15926 and 16384. The new class-wise random control uses NumPy's generator; the original subject-group allocation uses the released Python randomization routine. Identical seed numbers do not imply identical randomization algorithms across these two tasks.
\clearpage
\section{Reporting checklist}
Table~S11 lists the information needed to interpret further results from this benchmark. It complements the broader reporting and appraisal guidance in TRIPOD+AI and PROBAST+AI \citep{collins2024tripodai,moons2025probastai}.
\begin{table}[H]
\centering
\caption{Items needed to interpret a further result from this pipeline.}
\small
\begin{tabularx}{\textwidth}{p{3.3cm}X}
\toprule
Item & Detail to report\\
\midrule
Allocation unit & Case, independent subject, recording, seizure block or window; handling of repeated subject identities.\\
Analysis status & Original or later-added analysis; preregistered or exploratory; which rules were fixed before outcome inspection.\\
Target access & No target data, unlabeled adaptation, labeled calibration or within-patient fitting.\\
Temporal separation & Source-sample overlap, shared event membership, recording order, boundary exclusions and actual partition sizes.\\
Cohort eligibility & Counts before and after exclusions, represented seizure identities and reasons for unusable partitions.\\
Aggregation & Pooled windows, within-subject AUC or another explicit unit; treatment of repeated test appearances.\\
Threshold provenance & Scores and labels used for selection, tie handling, validation target and actual test outcome.\\
Alarm policy & Decision cadence, causal preprocessing, smoothing, persistence, merging, refractory periods and measured monitoring time.\\
Uncertainty & Independent units, dependence among repeated splits and the interpretation of the reported variation.\\
Chance comparison & Temporal surrogate construction and retained signal properties, where a formal chance claim is made.\\
External scope & Population, recording system, montage, preprocessing and which models were actually transferred.\\
Reproducibility & Dataset and code versions, manifests, prediction arrays and figure source tables.\\
\bottomrule
\end{tabularx}
\end{table}
\small
\printbibliography
\end{document}
