## Supplementary material for "Same pipelines, different answers: how evaluation design shapes EEG seizure prediction": Reproducibility archive (code, manifests and predictions): blocked.tex

\begin{table}[H]
\centering
\caption{PSD+LDA chronological block control: partition counts and case-level outcomes.}
\footnotesize
\begin{tabular}{lrrrrrr}
\toprule
Case & Train (positive) & Val (positive) & Test (positive) & Block AUC & Random AUC & $\Delta$AUC \\
\midrule
chb01 & 216 (148) & 235 (147) & 318 (148) & 0.724 & 0.848 & 0.124 \\
chb04 & 388 (148) & 613 (148) & 1183 (136) & 0.337 & 0.905 & 0.569 \\
chb05 & 212 (146) & 196 (146) & 336 (148) & 0.720 & 0.838 & 0.118 \\
chb06 & 978 (581) & 286 (148) & 180 (148) & 0.577 & 0.756 & 0.179 \\
chb08 & 176 (148) & 182 (148) & 228 (148) & 0.610 & 0.718 & 0.108 \\
chb09 & 423 (148) & 357 (148) & 408 (148) & 0.551 & 0.863 & 0.312 \\
chb10 & 701 (296) & 329 (296) & 274 (255) & 0.766 & 0.859 & 0.093 \\
chb13 & 366 (148) & 219 (146) & 20 (14) & 0.286 & 0.886 & 0.601 \\
chb14 & 339 (296) & 188 (146) & 252 (148) & 0.585 & 0.610 & 0.024 \\
chb15 & 577 (442) & 166 (148) & 169 (147) & 0.378 & 0.794 & 0.416 \\
chb22 & 350 (148) & 179 (148) & 131 (95) & 0.756 & 0.889 & 0.133 \\
\bottomrule
\end{tabular}
\par\smallskip{\footnotesize Random values average 20 allocations per case with identical partition class counts. $\Delta$AUC is random minus blocked. There were 11,175 retained windows, zero purged windows and no cross-partition block or raw-sample overlaps. Training fractions ranged from 17.8\% to 67.7\%; complete-block constraints took priority over nominal 60/20/20 window fractions.}
\end{table}
