## Supplementary material for "Same pipelines, different answers: how evaluation design shapes EEG seizure prediction": Reproducibility archive (code, manifests and predictions): boundaries.tex

\begin{table}[H]
\centering
\caption{Boundary audit of the original ten-case index allocation.}
\small
\begin{tabular}{lrrrr}
\toprule
Case & Index AUC & Time-sorted AUC & Shared event IDs & Overlapping pairs \\
\midrule
chb01 & 0.770 & 0.770 & 1 & 1 \\
chb03 & 0.836 & 0.836 & 1 & 1 \\
chb06 & 0.648 & 0.648 & 2 & 2 \\
chb07 & 0.407 & 0.407 & 1 & 1 \\
chb08 & 0.499 & 0.499 & 2 & 2 \\
chb10 & 0.939 & 0.939 & 1 & 2 \\
chb13 & 0.301 & 0.301 & 3 & 2 \\
chb14 & 0.478 & 0.478 & 2 & 2 \\
chb15 & 0.420 & 0.420 & 2 & 2 \\
chb22 & 0.776 & 0.776 & 1 & 1 \\
\bottomrule
\end{tabular}
\par\smallskip{\footnotesize AUC columns use PSD+LDA. Shared event IDs count distinct annotated seizures represented in more than one partition. Overlapping pairs count windows from the same EDF that share at least one source sample across partitions. A time reversal in chb03 did not change partition membership or AUC.}
\end{table}
