## Supplementary material for "Same pipelines, different answers: how evaluation design shapes EEG seizure prediction": Reproducibility archive (code, manifests and predictions): frequency.tex

\begin{table}[H]
\centering
\caption{Test appearances across 20 fixed subject-group partitions.}
\small
\begin{tabular}{lrlr}
\toprule
Subject group & Test $n$ & Subject group & Test $n$ \\
\midrule
chb01/chb21 & 4 & chb12 & 2 \\
chb02 & 1 & chb13 & 4 \\
chb03 & 3 & chb14 & 4 \\
chb04 & 2 & chb15 & 3 \\
chb05 & 5 & chb16 & 8 \\
chb06 & 1 & chb17 & 6 \\
chb07 & 4 & chb18 & 3 \\
chb08 & 6 & chb19 & 5 \\
chb09 & 1 & chb20 & 3 \\
chb10 & 2 & chb22 & 5 \\
chb11 & 5 & chb23 & 3 \\
\bottomrule
\end{tabular}
\par\smallskip{\footnotesize The 80 assignments equal four test groups per partition. chb01/chb21 is one independent group.}
\end{table}
