## Supplementary material for "Same pipelines, different answers: how evaluation design shapes EEG seizure prediction": Reproducibility archive (code, manifests and predictions): thresholds.tex

\begin{table}[H]
\centering
\caption{Validation-selected low-FPR thresholds on held-out subject groups.}
\small
\begin{tabular}{lccrc}
\toprule
Pipeline & Test FPR & Window sensitivity & Test $>1/60$ & Score discrepancy \\
\midrule
PSD+LDA & $0.0285\pm0.0351$ & $0.0299\pm0.0353$ & 10/20 & 4.8e-05 \\
EEGNet & $0.0181\pm0.0243$ & $0.0233\pm0.0311$ & 6/20 & 0.00035 \\
TCN & $0.0211\pm0.0186$ & $0.0324\pm0.0353$ & 9/20 & 0.00064 \\
1D-CNN & $0.0313\pm0.0371$ & $0.0556\pm0.0792$ & 10/20 & 0.00044 \\
EEG-Conformer & $0.0192\pm0.0257$ & $0.0243\pm0.0495$ & 6/20 & 5.1e-06 \\
\bottomrule
\end{tabular}
\par\smallskip{\footnotesize Values are mean $\pm$ sample SD across 20 dependent partitions. Every validation partition met the FPR target; achieved validation FPR was $0.01619\pm0.00032$ for each pipeline. Score discrepancy is the maximum absolute difference between recovered and archived test probabilities.}
\end{table}
