## Supplementary material for "Same pipelines, different answers: how evaluation design shapes EEG seizure prediction": Reproducibility archive (code, manifests and predictions): work_F_exclude_chb19_sensitivity.py

```
"""Drop-one sensitivity analysis for the chb19 case.

Uses existing per-patient 20-seed mean metrics and reports patient-weighted
summaries before and after removing chb19.
"""

from __future__ import annotations

from pathlib import Path

import numpy as np
import pandas as pd
from scipy.stats import mannwhitneyu


RESULTS_ROOT = Path(r"D:\seizure_results")
INPUT_CSV = RESULTS_ROOT / "per_patient_metrics.csv"
OUT_DIR = RESULTS_ROOT / "analysis_outputs" / "work_F_exclude_chb19"

MODELS = ["PSD+LDA", "EEGNet", "TCN", "1D-CNN", "EEG-Conformer"]

FOCAL = {
    "chb01",
    "chb02",
    "chb03",
    "chb04",
    "chb05",
    "chb07",
    "chb08",
    "chb10",
    "chb11",
    "chb15",
    "chb17",
    "chb22",
    "chb23",
}

NON_TARGET = {
    "chb06",
    "chb09",
    "chb12",
    "chb13",
    "chb14",
    "chb16",
    "chb18",
    "chb19",
    "chb20",
    "chb21",
}


def model_col(model: str, suffix: str) -> str:
    return f"{model}_{suffix}"


def iqr(values: np.ndarray) -> tuple[float, float, float]:
    values = values[~np.isnan(values)]
    if values.size == 0:
        return float("nan"), float("nan"), float("nan")
    return (
        float(np.nanmedian(values)),
        float(np.nanquantile(values, 0.25)),
        float(np.nanquantile(values, 0.75)),
    )


def cliffs_delta(x: np.ndarray, y: np.ndarray) -> float:
    x = x[~np.isnan(x)]
    y = y[~np.isnan(y)]
    if len(x) == 0 or len(y) == 0:
        return float("nan")
    greater = 0
    less = 0
    for xv in x:
        greater += int(np.sum(xv > y))
        less += int(np.sum(xv < y))
    return float((greater - less) / (len(x) * len(y)))


def add_group_labels(df: pd.DataFrame) -> pd.DataFrame:
    df = df.copy()

    def _group(pid: str) -> str:
        if pid in FOCAL:
            return "focal"
        if pid in NON_TARGET:
            return "non_target_localised"
        raise ValueError(f"Unknown patient ID: {pid}")

    df["onset_group"] = df["patient"].map(_group)
    return df


def model_means(df: pd.DataFrame) -> pd.DataFrame:
    rows = []
    df_no19 = df[df["patient"] != "chb19"]
    for model in MODELS:
        col = model_col(model, "auc")
        all_vals = df[col].to_numpy(float)
        no19_vals = df_no19[col].to_numpy(float)
        rows.append(
            {
                "model": model,
                "mean_auc_all_23": float(np.nanmean(all_vals)),
                "mean_auc_excluding_chb19": float(np.nanmean(no19_vals)),
                "delta_excl_minus_all": float(np.nanmean(no19_vals) - np.nanmean(all_vals)),
                "tractable_all_23_auc_gt_0p6": int(np.sum(all_vals > 0.6)),
                "tractable_excluding_chb19_auc_gt_0p6": int(np.sum(no19_vals > 0.6)),
                "n_all": int(np.sum(~np.isnan(all_vals))),
                "n_excluding_chb19": int(np.sum(~np.isnan(no19_vals))),
            }
        )
    return pd.DataFrame(rows)


def subgroup_table(df: pd.DataFrame, scenario: str) -> pd.DataFrame:
    rows = []
    for model in MODELS:
        col = model_col(model, "auc")
        focal = df.loc[df["onset_group"] == "focal", col].to_numpy(float)
        non_target = df.loc[df["onset_group"] == "non_target_localised", col].to_numpy(float)
        focal_valid = focal[~np.isnan(focal)]
        non_valid = non_target[~np.isnan(non_target)]

        f_med, f_q1, f_q3 = iqr(focal)
        n_med, n_q1, n_q3 = iqr(non_target)

        if len(focal_valid) > 0 and len(non_valid) > 0:
            _, p = mannwhitneyu(focal_valid, non_valid, alternative="two-sided")
        else:
            p = float("nan")

        rows.append(
            {
                "scenario": scenario,
                "model": model,
                "focal_n": int(len(focal_valid)),
                "non_target_n": int(len(non_valid)),
                "focal_median": f_med,
                "focal_q1": f_q1,
                "focal_q3": f_q3,
                "non_target_median": n_med,
                "non_target_q1": n_q1,
                "non_target_q3": n_q3,
                "delta_median_focal_minus_non_target": float(f_med - n_med),
                "focal_tractable_auc_gt_0p6": int(np.sum(focal_valid > 0.6)),
                "non_target_tractable_auc_gt_0p6": int(np.sum(non_valid > 0.6)),
                "cliffs_delta": cliffs_delta(focal, non_target),
                "mannwhitney_p": float(p),
            }
        )
    return pd.DataFrame(rows)


def write_report(means: pd.DataFrame, subgroup: pd.DataFrame) -> None:
    lines = []
    lines.append("# chb19 Drop-One Sensitivity")
    lines.append("")
    lines.append(
        "This analysis drops chb19 from the existing per-patient 20-seed mean "
        "metrics. It is a patient-weighted sensitivity analysis of the heatmap "
        "and subgroup summaries, not a retrained or seed-weighted PI-table "
        "analysis."
    )
    lines.append("")
    lines.append("## Model Means")
    lines.append("")
    lines.append(means.round(4).to_string(index=False))
    lines.append("")
    lines.append("## Subgroup Tests")
    lines.append("")
    lines.append(subgroup.round(4).to_string(index=False))
    lines.append("")
    lines.append(
        "Interpretation: if the deltas remain small after removing chb19, the "
        "artefact case is not driving the patient-weighted model means or the "
        "focal-vs-non-target-localised subgroup comparison."
    )
    (OUT_DIR / "exclude_chb19_report.md").write_text("\n".join(lines), encoding="utf-8")


def main() -> None:
    OUT_DIR.mkdir(parents=True, exist_ok=True)
    df = pd.read_csv(INPUT_CSV)
    df = add_group_labels(df)

    if set(df["patient"]) != FOCAL | NON_TARGET:
        missing = (FOCAL | NON_TARGET) - set(df["patient"])
        extra = set(df["patient"]) - (FOCAL | NON_TARGET)
        raise ValueError(f"Patient mapping mismatch. Missing={missing}, extra={extra}")

    means = model_means(df)
    subgroup_all = subgroup_table(df, "all_23")
    subgroup_no19 = subgroup_table(df[df["patient"] != "chb19"], "excluding_chb19")
    subgroup = pd.concat([subgroup_all, subgroup_no19], ignore_index=True)

    means_path = OUT_DIR / "TAB_model_means_drop_chb19.csv"
    subgroup_path = OUT_DIR / "TAB_subgroup_drop_chb19.csv"
    means.to_csv(means_path, index=False)
    subgroup.to_csv(subgroup_path, index=False)
    write_report(means, subgroup)

    print(f"Saved: {means_path}")
    print(f"Saved: {subgroup_path}")
    print(f"Saved: {OUT_DIR / 'exclude_chb19_report.md'}")
    print(means.round(4).to_string(index=False))


if __name__ == "__main__":
    main()
```
