## Supplementary material for "Same pipelines, different answers: how evaluation design shapes EEG seizure prediction": Reproducibility archive (code, manifests and predictions): work_G_ps_leakage_audit.py

```
"""Patient-specific split audit.

Compares chronological window splits with stratified random window splits on
the same patient set. The default run evaluates PSD+LDA only.
"""

from __future__ import annotations

import argparse
import json
import os
import random
from pathlib import Path

import numpy as np
import pandas as pd
import torch
import torch.nn as nn
from scipy.signal import welch
from scipy.stats import wilcoxon
from sklearn.discriminant_analysis import LinearDiscriminantAnalysis
from sklearn.model_selection import train_test_split

from data_utils import DATA_DIR, VALID_PATIENTS
from eval_utils import find_youden_threshold, full_evaluate
from models import CNN1D, EEGConformer, EEGNet, TCN


RESULTS_ROOT = Path(r"D:\seizure_results")
DEFAULT_OUT_DIR = RESULTS_ROOT / "analysis_outputs" / "work_G_ps_leakage"

FS = 256
BANDS = [(0.5, 4), (4, 8), (8, 13), (13, 30), (30, 40)]
DEVICE = torch.device("cuda" if torch.cuda.is_available() else "cpu")

MODEL_CFG = {
    "psd_lda": {"label": "PSD+LDA"},
    "1dcnn": {"label": "1D-CNN", "cls": CNN1D, "lr": 1e-4, "batch_size": 128, "dropout": None},
    "eegnet": {"label": "EEGNet", "cls": EEGNet, "lr": 1e-3, "batch_size": 128, "dropout": None},
    "tcn": {"label": "TCN", "cls": TCN, "lr": 1e-3, "batch_size": 128, "dropout": None},
    "eeg_conformer": {"label": "EEG-Conformer", "cls": EEGConformer, "lr": 1e-4, "batch_size": 64, "dropout": None},
}

MODEL_ALIASES = {
    "psd": "psd_lda",
    "lda": "psd_lda",
    "psd+lda": "psd_lda",
    "cnn": "1dcnn",
    "1d-cnn": "1dcnn",
    "conformer": "eeg_conformer",
    "eeg-conformer": "eeg_conformer",
}


def seed_everything(seed: int) -> None:
    random.seed(seed)
    np.random.seed(seed)
    torch.manual_seed(seed)
    if torch.cuda.is_available():
        torch.cuda.manual_seed_all(seed)


def stable_patient_offset(patient: str) -> int:
    return sum((i + 1) * ord(ch) for i, ch in enumerate(patient))


def load_patient(patient: str) -> tuple[np.ndarray, np.ndarray]:
    x_path = os.path.join(DATA_DIR, f"{patient}_X.npy")
    y_path = os.path.join(DATA_DIR, f"{patient}_y.npy")
    return np.load(x_path), np.load(y_path).astype(np.int64)


def has_two_classes(y: np.ndarray) -> bool:
    return len(np.unique(y)) == 2


def chronological_indices(y: np.ndarray) -> tuple[np.ndarray, np.ndarray, np.ndarray] | None:
    n = len(y)
    train_end = int(n * 0.6)
    val_end = int(n * 0.8)
    train_idx = np.arange(0, train_end)
    val_idx = np.arange(train_end, val_end)
    test_idx = np.arange(val_end, n)
    if not all(has_two_classes(y[idx]) for idx in (train_idx, val_idx, test_idx)):
        return None
    return train_idx, val_idx, test_idx


def random_indices(y: np.ndarray, seed: int) -> tuple[np.ndarray, np.ndarray, np.ndarray] | None:
    idx = np.arange(len(y))
    try:
        train_idx, temp_idx = train_test_split(
            idx,
            test_size=0.4,
            random_state=seed,
            stratify=y,
        )
        val_idx, test_idx = train_test_split(
            temp_idx,
            test_size=0.5,
            random_state=seed + 100_003,
            stratify=y[temp_idx],
        )
    except ValueError:
        return None
    return np.sort(train_idx), np.sort(val_idx), np.sort(test_idx)


def preictal_run_audit(y: np.ndarray) -> dict:
    split = chronological_indices(y)
    if split is None:
        return {"n_runs": np.nan, "n_boundary_split_runs": np.nan, "split_runs": ""}

    _, _, _ = split
    n = len(y)
    train_end = int(n * 0.6)
    val_end = int(n * 0.8)

    is_pre = (y == 1).astype(int)
    diff = np.diff(is_pre, prepend=0, append=0)
    starts = np.where(diff == 1)[0]
    ends = np.where(diff == -1)[0]

    split_runs = []
    for start, end in zip(starts, ends):
        touched = []
        if start < train_end and end > 0:
            touched.append("train")
        if start < val_end and end > train_end:
            touched.append("val")
        if end > val_end:
            touched.append("test")
        if len(touched) > 1:
            split_runs.append(f"{int(start)}:{int(end)}:{'/'.join(touched)}")

    return {
        "n_runs": int(len(starts)),
        "n_boundary_split_runs": int(len(split_runs)),
        "split_runs": ";".join(split_runs),
    }


def extract_psd(X: np.ndarray, chunk_size: int = 256) -> np.ndarray:
    chunks = []
    for start in range(0, len(X), chunk_size):
        chunk = X[start : start + chunk_size]
        freqs, pxx = welch(chunk, fs=FS, axis=-1, nperseg=512)
        feats = [
            pxx[:, :, (freqs >= lo) & (freqs <= hi)].mean(axis=-1)
            for lo, hi in BANDS
        ]
        chunks.append(np.concatenate(feats, axis=1).astype(np.float32))
    return np.vstack(chunks)


def run_psd_lda_features(
    X_psd: np.ndarray,
    y: np.ndarray,
    split: tuple[np.ndarray, np.ndarray, np.ndarray],
) -> dict:
    train_idx, val_idx, test_idx = split
    clf = LinearDiscriminantAnalysis(solver="svd", priors=[0.5, 0.5])
    clf.fit(X_psd[train_idx], y[train_idx])
    val_prob = clf.predict_proba(X_psd[val_idx])[:, 1]
    threshold = find_youden_threshold(y[val_idx], val_prob)
    test_prob = clf.predict_proba(X_psd[test_idx])[:, 1]
    return full_evaluate(y[test_idx], test_prob, threshold, stride_s=300)


class ArrayDataset(torch.utils.data.Dataset):
    def __init__(self, X: np.ndarray, y: np.ndarray):
        self.X = X
        self.y = y

    def __len__(self) -> int:
        return len(self.y)

    def __getitem__(self, idx: int):
        return torch.tensor(self.X[idx], dtype=torch.float32), torch.tensor(int(self.y[idx]), dtype=torch.long)


def make_loader(X: np.ndarray, y: np.ndarray, batch_size: int, weighted: bool, seed: int):
    dataset = ArrayDataset(X, y)
    if weighted:
        n0, n1 = int((y == 0).sum()), int((y == 1).sum())
        if n0 == 0 or n1 == 0:
            return None
        weights = np.where(y == 1, n0 / n1, 1.0).astype(np.float32)
        generator = torch.Generator()
        generator.manual_seed(seed)
        sampler = torch.utils.data.WeightedRandomSampler(
            torch.from_numpy(weights),
            num_samples=len(weights),
            replacement=True,
            generator=generator,
        )
        return torch.utils.data.DataLoader(dataset, batch_size=batch_size, sampler=sampler, num_workers=0)
    return torch.utils.data.DataLoader(dataset, batch_size=batch_size, shuffle=False, num_workers=0)


@torch.no_grad()
def collect_probs(model, loader) -> tuple[np.ndarray, np.ndarray]:
    model.eval()
    probs, labels = [], []
    for x, y in loader:
        logits = model(x.to(DEVICE))
        probs.append(torch.softmax(logits, dim=1)[:, 1].cpu().numpy())
        labels.append(y.numpy())
    return np.concatenate(probs), np.concatenate(labels)


def run_deep(
    model_key: str,
    X: np.ndarray,
    y: np.ndarray,
    split: tuple[np.ndarray, np.ndarray, np.ndarray],
    seed: int,
    max_epochs: int,
    patience: int,
) -> dict:
    cfg = MODEL_CFG[model_key]
    seed_everything(seed)
    train_idx, val_idx, test_idx = split

    train_loader = make_loader(X[train_idx], y[train_idx], cfg["batch_size"], weighted=True, seed=seed)
    val_loader = make_loader(X[val_idx], y[val_idx], cfg["batch_size"], weighted=False, seed=seed)
    test_loader = make_loader(X[test_idx], y[test_idx], cfg["batch_size"], weighted=False, seed=seed)
    if train_loader is None:
        raise ValueError("single-class training split")

    model = cfg["cls"]().to(DEVICE)
    optimizer = torch.optim.AdamW(model.parameters(), lr=cfg["lr"], weight_decay=1e-4)
    scheduler = torch.optim.lr_scheduler.ReduceLROnPlateau(
        optimizer, mode="max", factor=0.5, patience=5, min_lr=1e-7
    )
    criterion = nn.CrossEntropyLoss()

    best_auc = -np.inf
    best_state = {k: v.detach().cpu().clone() for k, v in model.state_dict().items()}
    patience_left = patience

    for _ in range(max_epochs):
        model.train()
        for bx, by in train_loader:
            bx, by = bx.to(DEVICE), by.to(DEVICE)
            optimizer.zero_grad()
            loss = criterion(model(bx), by)
            loss.backward()
            nn.utils.clip_grad_norm_(model.parameters(), 1.0)
            optimizer.step()

        val_prob, val_y = collect_probs(model, val_loader)
        try:
            val_auc = float(full_evaluate(val_y, val_prob, 0.5)["auc"])
        except ValueError:
            val_auc = 0.5
        scheduler.step(val_auc)

        if val_auc > best_auc:
            best_auc = val_auc
            best_state = {k: v.detach().cpu().clone() for k, v in model.state_dict().items()}
            patience_left = patience
        else:
            patience_left -= 1
            if patience_left <= 0:
                break

    model.load_state_dict(best_state)
    model.to(DEVICE)
    val_prob, val_y = collect_probs(model, val_loader)
    threshold = find_youden_threshold(val_y, val_prob)
    test_prob, test_y = collect_probs(model, test_loader)
    return full_evaluate(test_y, test_prob, threshold, stride_s=300)


def normalise_models(models: list[str]) -> list[str]:
    if len(models) == 1 and models[0].lower() == "all":
        return list(MODEL_CFG)
    keys = []
    for model in models:
        key = MODEL_ALIASES.get(model.lower(), model.lower())
        if key not in MODEL_CFG:
            raise ValueError(f"Unknown model: {model}")
        keys.append(key)
    return keys


def metric_row(patient: str, model_key: str, mode: str, repeat: int, metrics: dict) -> dict:
    return {
        "patient": patient,
        "model_key": model_key,
        "model": MODEL_CFG[model_key]["label"],
        "split_mode": mode,
        "repeat": repeat,
        "auc": metrics["auc"],
        "sensitivity": metrics["sensitivity"],
        "specificity": metrics["specificity"],
        "precision": metrics["precision"],
        "f1": metrics["f1"],
        "far": metrics["far"],
        "event_sensitivity": metrics["event_sensitivity"],
        "n_events": metrics["n_events"],
        "threshold": metrics["threshold"],
    }


def paired_summary(results: pd.DataFrame) -> pd.DataFrame:
    rows = []
    random_mean = (
        results[results["split_mode"] == "random_window"]
        .groupby(["patient", "model_key", "model"], as_index=False)["auc"]
        .mean()
        .rename(columns={"auc": "random_auc_mean"})
    )
    chrono = results[results["split_mode"] == "chronological_window"][
        ["patient", "model_key", "model", "auc"]
    ].rename(columns={"auc": "chronological_auc"})
    paired = chrono.merge(random_mean, on=["patient", "model_key", "model"], how="inner")
    paired["delta_random_minus_chronological"] = paired["random_auc_mean"] - paired["chronological_auc"]

    for (model_key, model), sub in paired.groupby(["model_key", "model"]):
        delta = sub["delta_random_minus_chronological"].to_numpy(float)
        if len(delta) >= 2 and np.any(delta != 0):
            try:
                _, p = wilcoxon(delta, alternative="two-sided")
            except ValueError:
                p = np.nan
        else:
            p = np.nan
        rows.append(
            {
                "model_key": model_key,
                "model": model,
                "n_patients": len(sub),
                "chronological_auc_mean": float(sub["chronological_auc"].mean()),
                "random_auc_mean": float(sub["random_auc_mean"].mean()),
                "delta_mean": float(delta.mean()),
                "delta_median": float(np.median(delta)),
                "delta_sd": float(delta.std(ddof=1)) if len(delta) > 1 else np.nan,
                "wilcoxon_p": float(p) if not np.isnan(p) else np.nan,
            }
        )
    return pd.DataFrame(rows), paired


def main() -> None:
    parser = argparse.ArgumentParser()
    parser.add_argument("--models", nargs="+", default=["psd_lda"])
    parser.add_argument("--patients", nargs="*", default=None)
    parser.add_argument("--n-random", type=int, default=20)
    parser.add_argument("--seed", type=int, default=20260517)
    parser.add_argument("--max-epochs", type=int, default=100)
    parser.add_argument("--patience", type=int, default=20)
    parser.add_argument("--out-dir", type=Path, default=DEFAULT_OUT_DIR)
    args = parser.parse_args()

    out_dir = args.out_dir
    out_dir.mkdir(parents=True, exist_ok=True)
    models = normalise_models(args.models)
    candidate_patients = args.patients or VALID_PATIENTS

    rows = []
    audit_rows = []
    skipped = []

    for patient in candidate_patients:
        print(f"[patient] {patient}", flush=True)
        X, y = load_patient(patient)
        X_psd = extract_psd(X) if "psd_lda" in models else None
        chrono_split = chronological_indices(y)
        audit = preictal_run_audit(y)
        audit.update({"patient": patient, "n_windows": len(y), "n_preictal": int((y == 1).sum())})
        audit_rows.append(audit)

        if chrono_split is None:
            skipped.append({"patient": patient, "reason": "chronological split lacks both classes"})
            continue

        for model_key in models:
            try:
                print(f"  [{model_key}] chronological", flush=True)
                if model_key == "psd_lda":
                    metrics = run_psd_lda_features(X_psd, y, chrono_split)
                else:
                    metrics = run_deep(
                        model_key,
                        X,
                        y,
                        chrono_split,
                        seed=args.seed,
                        max_epochs=args.max_epochs,
                        patience=args.patience,
                    )
                rows.append(metric_row(patient, model_key, "chronological_window", 0, metrics))
            except Exception as exc:
                skipped.append({"patient": patient, "model_key": model_key, "reason": f"chrono failed: {exc}"})
                continue

            for repeat in range(args.n_random):
                print(f"  [{model_key}] random repeat {repeat + 1}/{args.n_random}", flush=True)
                split_seed = args.seed + repeat * 10_007 + stable_patient_offset(patient)
                rand_split = random_indices(y, split_seed)
                if rand_split is None:
                    skipped.append({"patient": patient, "model_key": model_key, "reason": "random split failed"})
                    continue
                try:
                    if model_key == "psd_lda":
                        metrics = run_psd_lda_features(X_psd, y, rand_split)
                    else:
                        metrics = run_deep(
                            model_key,
                            X,
                            y,
                            rand_split,
                            seed=split_seed,
                            max_epochs=args.max_epochs,
                            patience=args.patience,
                        )
                    rows.append(metric_row(patient, model_key, "random_window", repeat, metrics))
                except Exception as exc:
                    skipped.append({"patient": patient, "model_key": model_key, "reason": f"random failed: {exc}"})

    results = pd.DataFrame(rows)
    audit = pd.DataFrame(audit_rows)
    skipped_df = pd.DataFrame(skipped)
    summary, paired = paired_summary(results) if not results.empty else (pd.DataFrame(), pd.DataFrame())

    results.to_csv(out_dir / "ps_split_results.csv", index=False)
    paired.to_csv(out_dir / "ps_split_paired.csv", index=False)
    summary.to_csv(out_dir / "ps_split_summary.csv", index=False)
    audit.to_csv(out_dir / "chronological_boundary_audit.csv", index=False)
    skipped_df.to_csv(out_dir / "skipped.csv", index=False)

    report = {
        "models": models,
        "n_random": args.n_random,
        "n_result_rows": int(len(results)),
        "summary": summary.to_dict(orient="records"),
        "n_skipped": int(len(skipped_df)),
    }
    (out_dir / "ps_split_report.json").write_text(json.dumps(report, indent=2), encoding="utf-8")

    print(f"Saved outputs to {out_dir}")
    if not summary.empty:
        print(summary.to_string(index=False))
    if not skipped_df.empty:
        print(f"Skipped/failed rows: {len(skipped_df)}")


if __name__ == "__main__":
    main()
```
